# A Genetic Atlas of Relationships Between Circulating Metabolites and Liability to Psychiatric Conditions

**DOI:** 10.1101/2025.05.19.25327956

**Authors:** Dylan J. Kiltschewskij, William R. Reay, Murray J. Cairns

## Abstract

Circulating metabolites have been observed to be altered in psychiatric conditions and could be clinically actionable. To explore this question we used the largest genome-wide association studies available to investigate genetic correlation and causal relationships between 10 psychiatric conditions and 249 circulating metabolites. This revealed 1,100 trait pairings, involving fatty acids, lipoproteins and other metabolites, with evidence for causal effects on the liability for major depressive disorder, post-traumatic stress disorder and anorexia nervosa. Notably, the most robust association was a putative causal effect of high-density lipoprotein on anorexia nervosa. We also observed significant relatonships between metabolic traits and cortical thickness and surface area, as well as evidence of shared gene-level common variant associations amongst 23 metabolite-psychiatric pairings, converging in pathways with metabolic and neuronal function. These findings highlight specific metabolites as potential biomarkers and therapeutic targets in the clinical management of psychiatric disorders.

## INTRODUCTION

Psychiatric conditions, such as schizophrenia, major depressive disorder and bipolar disorder, are associated with a variety of comorbid diagnoses that impede effective clinical management, increase mortality and decrease quality of life [1–3]. Individuals with a psychiatric condition exhibit strong overrepresentation of cardiometabolic conditions, including metabolic syndrome and coronary artery disease, which contribute to an 80% higher risk of premature death due to heart disease and a 10-to-20-year shorter life expectancy [1–3]. Alarmingly, while improved lifestyle and healthcare in recent decades have reduced the cardiometabolic burden in the general population, the incidence and mortality has remained stubbornly high amongst individuals with psychiatric illness [4, 5]. This is likely a reflection of the complex relationship between cardiometabolic traits and psychiatric illness, which is generally thought to arise from a combination of psychotropic medications, poor lifestyle and systemic barriers to appropriate care [1–7]. Recent evidence suggests, however, that psychiatric and cardiometabolic traits share genetic components and causal relationships that make them targets for therapeutic intervention. This supports further investigation to capitalise on the potential therapeutic benefits for decreasing the burden of both the psychiatric symptoms and adverse cardiometabolic outcomes [8–14].

Large genome-wide association studies (GWAS) have uncovered strong common variant associations with psychiatric [15–24] and cardiometabolic traits [25, 26]. Recent advances in statistical genetics now provide the unprecedented opportunity to capitalise on the power of large GWAS to further explore shared biology in this comorbidity and prioritise interventions for clinical trials. For instance, GWAS-guided exploration of drug repurposing candidates suggests drug related to cardiometabolic health modify risk for some psychiatric conditions. Specifically, omega-3 nutraceuticals exhibit evidence for a beneficial effect in bipolar disorder, while antihypertensive angiotensin-converting enzyme (ACE) inhibitors may be risk increasing in schizophrenia [8]. We have also observed extensive genetic correlation amongst blood-based biomarkers, psychiatric illness and brain anatomy, with genetic causal inference further identifying causal relationships involving biomarkers such as glycaemic traits and C-reactive protein [9–11]. However, lipid-related traits – such as fatty acids, cholesterol and phospholipids – and other metabolites remain relatively underexplored in this context, despite their intrinsic role in neuronal function and emergence as a key feature of psychiatric conditions [27, 28]. Indeed, many of these traits impact biological processes central to the pathogenesis of psychiatric illness, exhibit dysregulation in these conditions, and, critically, can contribute to the development of cardiometabolic conditions [27, 28]. Although circulating metabolites represent an attractive prospect for preventative intervention, since many are clinically actionable through existing pharmacological and dietary interventions, the extent of shared genetic architecture and causal relationships between metabolites and psychiatric illness remains poorly characterised. To address this, we leveraged the largest publicly available GWAS of metabolites assayed using high-throughput metabolomics to comprehensively examine genetic correlation and causation between 249 circulating metabolites and 10 psychiatric conditions. Our analyses revealed extensive genetic correlation involving lipoproteins, phospholipids, triglycerides, fatty acids and other metabolic traits, while evidence for causal relationships was also uncovered. Furthermore, we report genes with common variant associations for both metabolites and psychiatric conditions that provide further insights into the interrelationship between these traits.

## RESULTS

### Extensive genetic correlation amongst circulating metabolites and psychiatric conditions

A schematic overview of the current study is presented in Figure 1. Genetic correlation was examined between 249 blood-based metabolites and 10 psychiatric conditions via linkage disequilibrium score regression (LDSR). This analysis utilised the largest publicly available and uniformly processed metabolite GWAS conducted by Tambets *et al.* [25], wherein common variant signatures for blood metabolites were meta-analysed using individuals of European ancestry from the UK (*N* = 413,897) and Estonian (*N* = 185,352) Biobanks. The metabolites include 81 lipid, amino acid, and glycolysis-related traits (e.g. total triglycerides, tyrosine and glucose), herein referred to as “primary metabolites” (See Table S1 for a full overview of all traits). The remaining traits relate to lipoprotein absolute lipid content (98 traits) or lipid ratios (70 traits), subcategorised by lipoprotein diameter (e.g. triglycerides in small, medium, large and very large high density lipoprotein (HDL); [25]).

**Figure 1.**
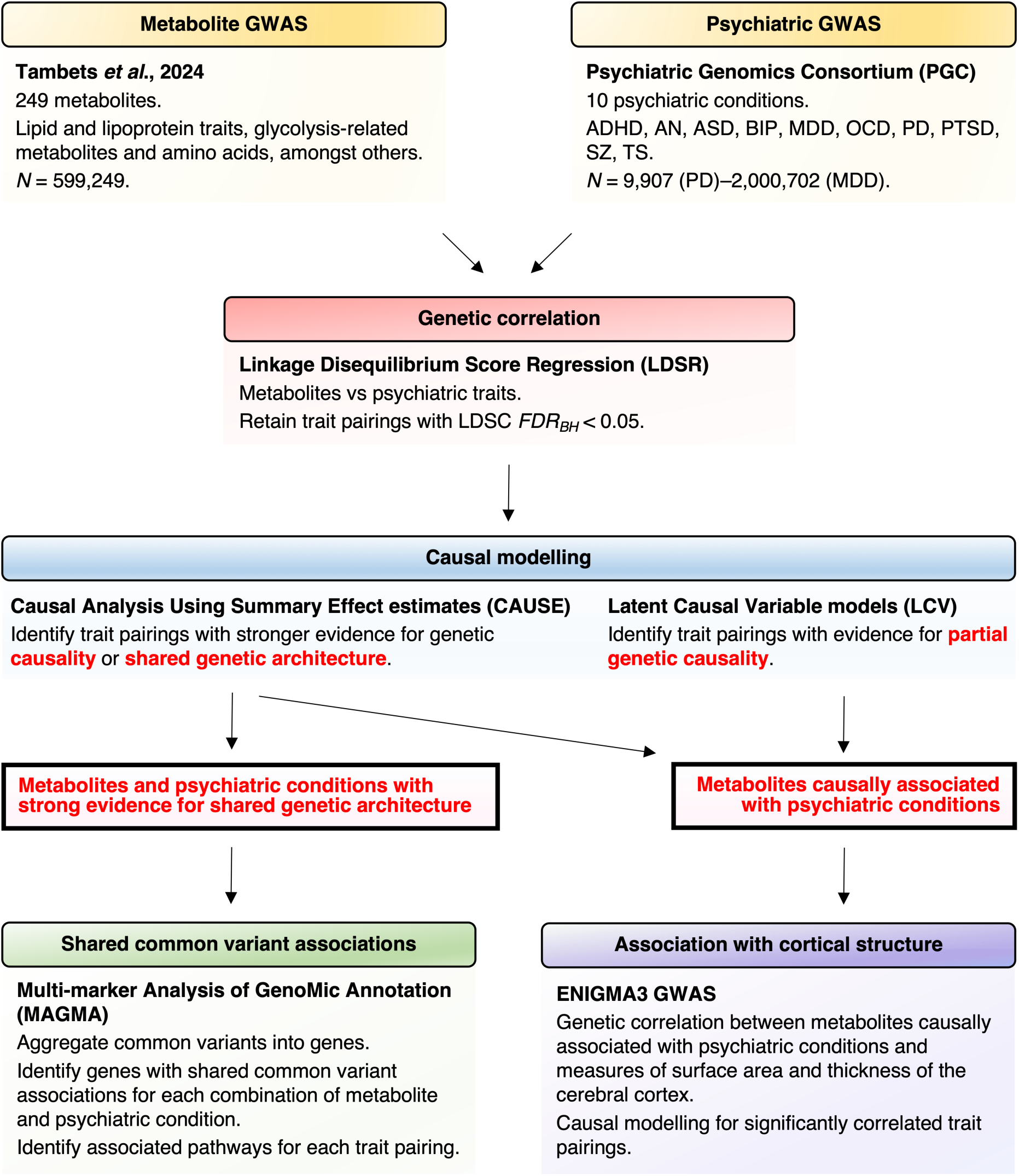
Schematic overview of study design. GWAS summary statistics for 249 circulating metabolites were obtained from a recent meta-analysis of ∼600,000 individuals from the Estonian and UK Biobanks [25]. Summary statistics for 10 psychiatric conditions were obtained from the Psychiatric Genomics Consortium. Genetic correlation was firstly examined between all pairings of metabolite and psychiatric condition via linkage disequilibrium score regression (LDSR) [29, 30]. A total of 1,100 significant trait pairings (*FDR_BH_* < 0.05) were subjected to causal inference using Latent Causal Variable (LCV) [31] and Causal Analysis Using Summary Effect estimates (CAUSE) models [32]. Metabolites with evidence for causality were further examined for genetic relationships with respect to measures of cortical structure using GWAS from the ENIGMA consortium. CAUSE also identified 23 trait pairings with stronger evidence for shared genetic architecture than a causal relationship. Overlapping biology was further explored between these traits by identifying shared gene and gene-set associations via the Multi-marker Analysis of GenoMic Annotation (MAGMA) [33].

Overall, 1,100 metabolite-psychiatric trait pairings surpassed correction for multiple testing (*FDR_BH_* < 0.05; Fig. 2a, Fig S1, Table S2). Interestingly, all psychiatric conditions except panic disorder (PD) were correlated with at least one metabolite, with particularly strong representation for attention-deficit/hyperactivity disorder (ADHD, 210 metabolites); anorexia nervosa (AN, 174), major depressive disorder (MDD, 199), obsessive compulsive disorder (OCD, 154), post-traumatic stress disorder (PTSD, 184) and schizophrenia (SZ, 134; Fig. 2b). Amongst the primary metabolites, fatty acid traits were most frequently correlated with psychiatric illness (92 correlations), followed by amino acids (31), triglycerides (24) and lipoprotein particle sizes (21; Fig. 2c, Table S2). In addition, we also observed extensive correlation of lipoprotein absolute lipid content (430 correlations) and lipid ratios (345) across a range of lipoprotein subclasses (Fig. S1, Table S2). The top three genetic correlations ranked by absolute *Z* scores are as follows: glycoprotein acetyls and MDD (*r_g_* = 0.2, *SE* = 0.02, *P* = 2.11 × 10^−38^), ratio of monounsaturated fatty acids to total fatty acids and MDD (*r_g_* = 0.22, *SE* = 0.02, *P* = 1.2 × 10^−34^) and degree of unsaturation and ADHD (*r_g_* = –0.3, *SE* = 0.02, *P* = 1.66 × 10^−35^; Table S2).

**Figure 2.**
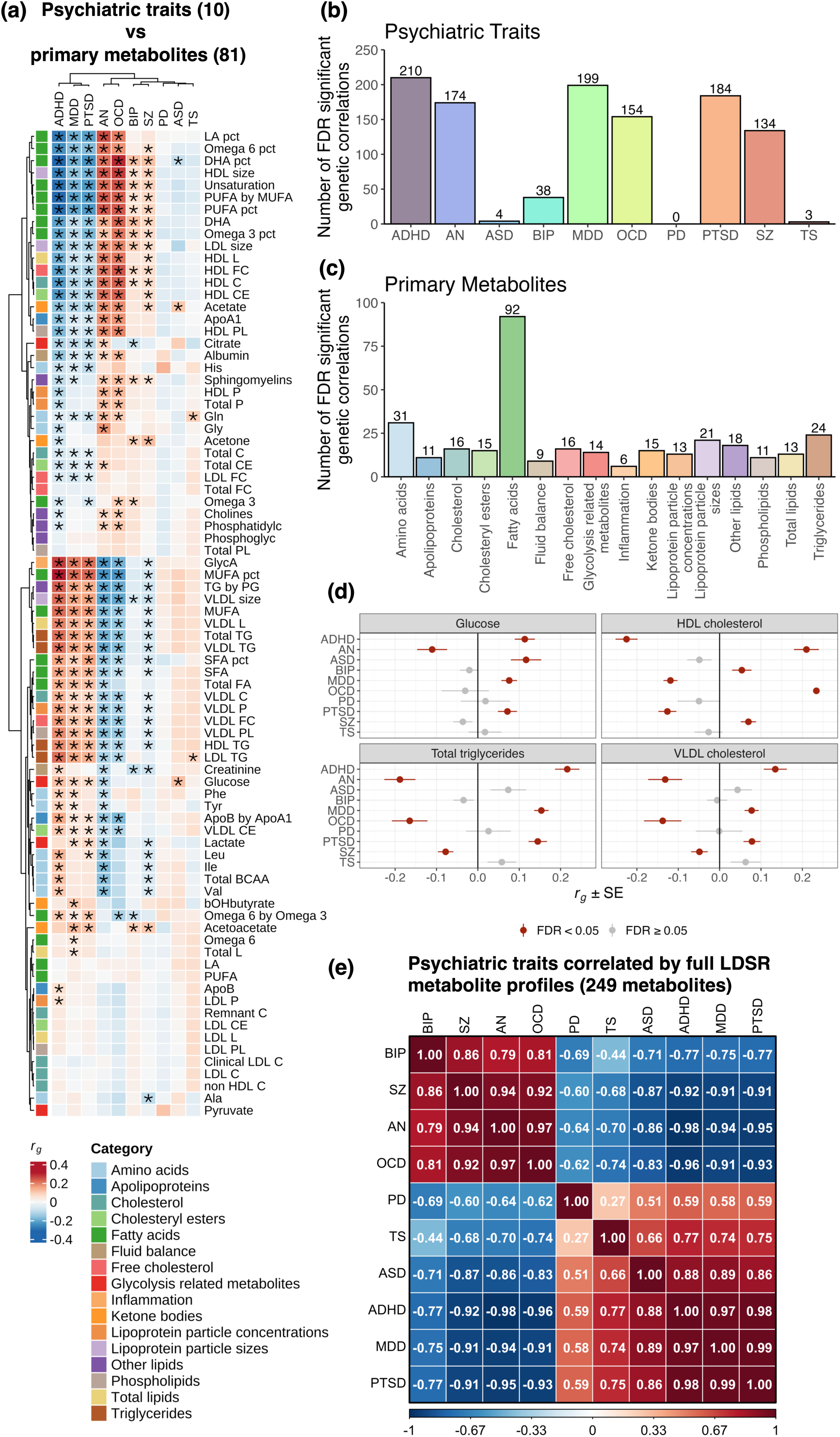
Genetic correlation amongst circulating metabolites and psychiatric traits. **(a)** Heatmap depicting LDSR genetic correlation coefficients (*r_g_*) between the 81 primary metabolites and 10 psychiatric conditions. Rows and columns were subject to hierarchal clustering to identify similar groups of traits. \**FDR_BH_* < 0.05. Full metabolite names can be accessed in Table S1, and data for the lipoprotein traits subcategorised by particle diameter can be accessed in Table S2 and Fig. S1. **(b)** The number of significant metabolite correlations (including lipoprotein traits subcategorised by particle diameter) per psychiatric trait. **(c)** The number of significant psychiatric correlations for the 81 primary metabolites. **(d)** Forest plots depicting key examples of metabolic traits exhibiting divergent patterns of genetic correlation with respect to psychiatric traits. Data plotted as *r_g_* ± standard error (*SE*). **(e)** Heatmap of Pearson correlation amongst psychiatric traits upon pairwise comparison of metabolite LDSR genetic correlation profiles (transformed into *Z*-scores [i.e. *r_g_* / *SE*]). Rows and columns were subject to hierarchal clustering to identify psychiatric conditions with the most similar metabolite correlation profiles.

For many metabolites, we consistently observed divergent patterns of genetic correlation (Fig. 2d). For instance, total triglycerides and very low-density lipoprotein (VLDL) cholesterol were positively correlated with ADHD, MDD and PTSD, and negatively correlated with AN, OCD and SZ (Fig. 2d). This divergence was further apparent upon comparison of metabolite LDSR profiles between the psychiatric conditions, noting that genetic correlations were transformed into *Z*-scores (i.e. *r_g_* / *SE*) to capture the magnitude and significance of each estimate (Fig. 2e, Table S3). Interestingly, AN, BIP, OCD and SZ exhibited strong positive correlation of metabolite LDSR profiles (Pearson *r* ≥ 0.79, *P* ≤ 2.36 × 10^−55^), and all four were negatively correlated with the remaining six psychiatric conditions (Pearson *r* ≤ –0.44, *P* ≤ 1.71 × 10^−13^). Among these six conditions, ADHD, MDD and PTSD exhibited particularly strong correlation of metabolite LDSR profiles (Pearson *r* ≥ 0.97, *P* ≤ 1.02 × 10^−157^). These results collectively suggest that psychiatric conditions share varying degrees of genetic correlation with circulating metabolites.

### Evidence for metabolites exhibiting partial genetic causality on psychiatric conditions

Partial genetic causality was examined between all significantly correlated metabolites and psychiatric conditions using latent causal variable (LCV) models [31]. These models specifically compare variant marginal effect size distributions between traits to determine whether one trait exhibits evidence for partial genetic causality on the other, expressed as a posterior genetic causality proportion (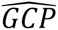). We note that these 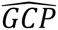 estimates quantify the strength of evidence that partial genetic causation exists, rather than the magnitude of causality. Two trait pairings exhibited strong evidence for partial genetic causality after correction for multiple testing (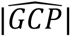 ≥ 0.6, *FDR_BH_* < 0.05): phospholipids to total lipids ratio in chylomicrons and extremely large VLDL → MDD (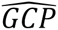 = 0.65) and cholesterol to total lipids ratio in medium low-density lipoprotein (LDL) → PTSD (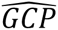 = 0.63; Table 1; Table S4). By integrating the corresponding LDSR genetic correlation coefficients, we can infer that elevation of these metabolites is respectively associated with increased risk for MDD (*r_g_* =0.16) and decreased for PTSD (*r_g_* = –0.19; Table S2). Using independent metabolite GWAS from [26], directionally consistent 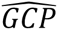 estimates were uncovered for these relationships (≥ 0.18; Table S5). While this did not exceed our criteria for strong evidence of partial genetic causality, we note these GWAS are of substantially reduced sample size (*N* = 136,016) compared to those used for the primary analysis (*N* = 599,529).

**Table 1.**
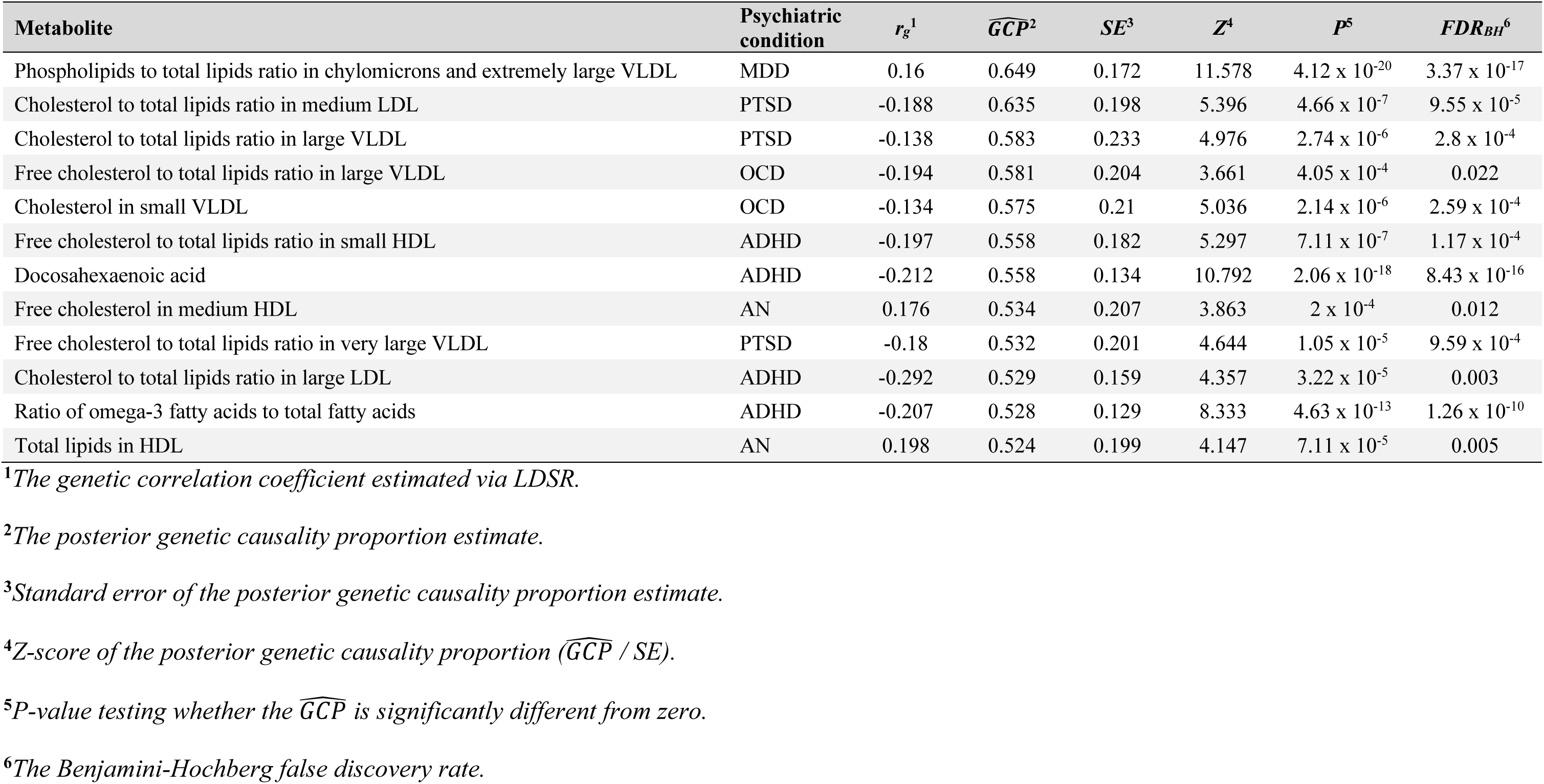
Metabolites and psychiatric conditions with strong or moderate evidence of partial genetic causality.

| Metabolite | Psychiatric condition | $r_g^1$ | $\widehat{GCP}^2$ | $SE^3$ | $Z^4$ | $P^5$ | $FDR_{BH}^6$ |
| --- | --- | --- | --- | --- | --- | --- | --- |
| Phospholipids to total lipids ratio in chylomicrons and extremely large VLDL | MDD | 0.16 | 0.649 | 0.172 | 11.578 | $4.12 \times 10^{-20}$ | $3.37 \times 10^{-17}$ |
| Cholesterol to total lipids ratio in medium LDL | PTSD | -0.188 | 0.635 | 0.198 | 5.396 | $4.66 \times 10^{-7}$ | $9.55 \times 10^{-5}$ |
| Cholesterol to total lipids ratio in large VLDL | PTSD | -0.138 | 0.583 | 0.233 | 4.976 | $2.74 \times 10^{-6}$ | $2.8 \times 10^{-4}$ |
| Free cholesterol to total lipids ratio in large VLDL | OCD | -0.194 | 0.581 | 0.204 | 3.661 | $4.05 \times 10^{-4}$ | 0.022 |
| Cholesterol in small VLDL | OCD | -0.134 | 0.575 | 0.21 | 5.036 | $2.14 \times 10^{-6}$ | $2.59 \times 10^{-4}$ |
| Free cholesterol to total lipids ratio in small HDL | ADHD | -0.197 | 0.558 | 0.182 | 5.297 | $7.11 \times 10^{-7}$ | $1.17 \times 10^{-4}$ |
| Docosahexaenoic acid | ADHD | -0.212 | 0.558 | 0.134 | 10.792 | $2.06 \times 10^{-18}$ | $8.43 \times 10^{-16}$ |
| Free cholesterol in medium HDL | AN | 0.176 | 0.534 | 0.207 | 3.863 | $2 \times 10^{-4}$ | 0.012 |
| Free cholesterol to total lipids ratio in very large VLDL | PTSD | -0.18 | 0.532 | 0.201 | 4.644 | $1.05 \times 10^{-5}$ | $9.59 \times 10^{-4}$ |
| Cholesterol to total lipids ratio in large LDL | ADHD | -0.292 | 0.529 | 0.159 | 4.357 | $3.22 \times 10^{-5}$ | 0.003 |
| Ratio of omega-3 fatty acids to total fatty acids | ADHD | -0.207 | 0.528 | 0.129 | 8.333 | $4.63 \times 10^{-13}$ | $1.26 \times 10^{-10}$ |
| Total lipids in HDL | AN | 0.198 | 0.524 | 0.199 | 4.147 | $7.11 \times 10^{-5}$ | 0.005 |

<sup>1</sup>The genetic correlation coefficient estimated via LDSR.
<sup>2</sup>The posterior genetic causality proportion estimate.
<sup>3</sup>Standard error of the posterior genetic causality proportion estimate.
<sup>4</sup>Z-score of the posterior genetic causality proportion ( $\widehat{GCP} / SE$ ).
<sup>5</sup>P-value testing whether the $\widehat{GCP}$ is significantly different from zero.
<sup>6</sup>The Benjamini-Hochberg false discovery rate.

A further 10 metabolites exhibited moderate evidence for partial genetic causality (0.5 ≤ 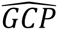 < 0.6, *FDR_BH_* < 0.05) on a psychiatric condition (Table 1, Table S4). Of these, four were associated with ADHD: docosahexaenoic acid (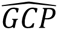 = 0.56), free cholesterol to total lipids ratio in small high-density lipoprotein (HDL; 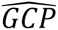 = 0.56), cholesterol to total lipids ratio in large LDL (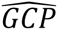 = 0.53), and ratio of omega-3 fatty acids to total fatty acids (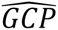 = 0.53). The remaining relationships included two HDL-related traits → AN, two VLDL-related traits → OCD and a further two VLDL-related traits → PTSD (Table 1, Table S4). Across all 10 trait pairings, the genetic correlation coefficients revealed that elevation of these metabolites is associated with decreased odds of their corresponding psychiatric conditions (*r_g_* ≤ –0.13), except for the HDL-related traits → AN, which were risk-increasing (*r_g_* ≥ 0.18; Table 1, Table S2). Interestingly, four of these relationships exhibited strong or moderate evidence for partial genetic causality using metabolite GWAS from [26], despite the decreased sample size: docosahexaenoic acid → ADHD (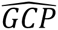 = 0.61), cholesterol to total lipids ratio in large VLDL → ADHD (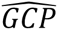 = 0.72), total lipids in HDL → AN (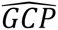 = 0.82) and free cholesterol to total lipids ratio in large VLDL → OCD (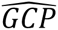 = 0.53; Table S5).

### Putative causal relationships uncovered for HDL-related traits on AN

We further explored evidence for causality amongst genetically correlated traits using the Causal Analysis Using Summary Effect estimates (CAUSE) model [32], which applies a Bayesian approach to Mendelian randomisation (MR). Specifically, CAUSE uses independent single nucleotide polymorphisms (SNPs) associated with an exposure trait (*Pgwas* < 1 × 10^−3^) to obtain a causal estimate of the exposure (i.e. metabolites) on an outcome trait (i.e. psychiatric conditions). These estimates are conditioned on horizontal pleiotropy (i.e. a variant acting on two traits through separate pathways), modelled using a prior beta distribution that estimates the proportion of variants associated with horizontal pleiotropy rather than a causal relationship. We specifically compared models of causality and horizontal pleiotropy together (“causal”) versus models of horizontal pleiotropy only (“sharing”). This is expressed as a change in expected log pointwise posterior density (*ΔELPD*), wherein negative values indicate the causal model fits better than the sharing.

Using prior distributions that assume moderate (q ∼ beta(1,10)) or low (q ∼ beta(1,10)) horizontal pleiotropy, five traits related to HDL exhibited stronger evidence for causality than horizontal pleiotropy with respect to AN (*FDR_BH_* < 0.05, ΔELPD < 0; Fig. 3a, 3b, Table S6). These include: average diameter for HDL particles, cholesterol/cholesteryl esters in very large HDL, concentration of large HDL particles, and free cholesterol to total lipids ratio in very large HDL. Although these trait pairings did not survive correction for multiple testing using the most conservative prior distribution (q ∼ beta(1,2)), all ΔELPD estimates were directionally consistent (Fig. 3b, Table S6). Reverse analyses utilising AN as the exposure trait also revealed no evidence for reverse causality, suggesting there is no bidirectional relationship between these traits (Fig. S2, Table S7). In contrast, MDD (six metabolites) and PTSD (two metabolites) exhibited evidence for causal relationships in the forwards analysis but also displayed evidence for reverse causality, and thus were not analysed further (Fig. 3a, Fig. S2, Table S6, S7).

**Figure 3.**
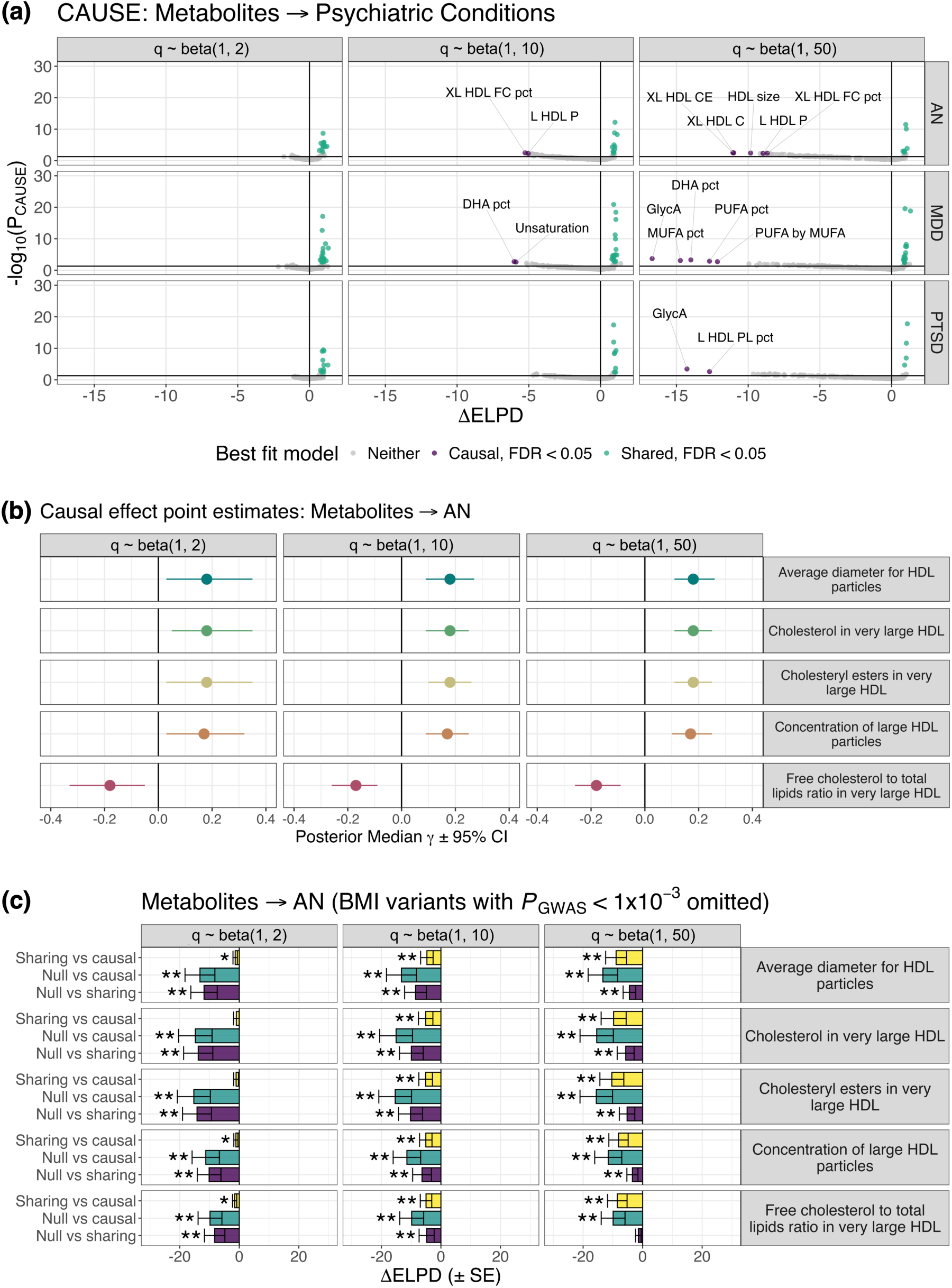
Causal modelling between metabolites and psychiatric illness using CAUSE. **(a)** Scatter plots summarising CAUSE models examining metabolites with evidence for a causal relationship with anorexia nervosa (AN), major depressive disorder (MDD) or post-traumatic stress disorder (PTSD). Each point represents a pairing of metabolite and psychiatric condition, noting these results are restricted to psychiatric conditions with evidence for at least one causal relationship. The x-axis corresponds to the change in expected log pointwise posterior density (*ΔELPD*) for CAUSE sharing and causal models (x-axis), with more negative values indicating causal models fit better than sharing models, and *vice versa*. The corresponding –log_10_(*P*) values are plotted on the left y-axis. Purple points = trait pairings with stronger evidence for causality at an *FDR_BH_* < 0.05; green points = trait pairings with stronger evidence for horizontal pleiotropy at an *FDR_BH_* < 0.05. Horizontal black line corresponds to a nominal *P* < 0.05. Facet headings indicate parameters used to define the prior beta distributions, with alpha = 1, beta = 2 representing the most conservative prior, and alpha =1, beta = 50 the most relaxed. Full results for all genetically correlated trait pairings identified via LDSR are available in Table S6. **(b)** Causal effect estimates (posterior median γ ± 95% credible interval [*CI*]) summarising direction of effect between all five HDL-related traits causally associated with AN. **(c)** *ΔELPD* (± standard error [*SE*]) estimates for the five HDL-related traits on AN after excluding variants associated with BMI at a *P_GWAS_* < 1 × 10^−3^ [35]. Yellow bars = the sharing versus causal model comparisons, similar to those depicted in panel **(a)**; teal bars = comparison of casual and null models; purple bars = comparison of sharing and null models. Note that a negative *ΔELPD* estimate indicates that the second model (as indicated on the left y-axis) fits better than the first. * = *P* < 0.05, ** = *FDR_BH_* < 0.05.

For HDL-related traits → AN, the sign of the causal effect term (posterior median γ) was generally positive, indicating that elevation of each HDL trait was associated with increased risk for AN (Fig. 3b, Table S6). The exception was free cholesterol to total lipids ratio in very large HDL, which was associated with decreased AN risk (Fig. 3b, Table S6). The AN GWAS was composed primarily of cohorts diagnosed using DSM-IV, which utilises low BMI as a criterion for AN diagnosis, thereby potentially confounding the relationship between HDL and AN, as reviewed elsewhere [34]. Thus, these analyses were then repeated after excluding all SNPs associated with BMI at a *P_GWAS_* < 1 × 10^−3^, using GWAS summary statistics from [35].

Strikingly, all causal relationships remained significant and directionally consistent (Fig, 3c, Fig. S3, Table S8). However, no evidence for causality was observed for the forwards analysis when utilising metabolite GWAS from [26] (Table S9). This is likely since CAUSE is more sensitive to smaller sample sizes in these GWAS than LCV, as SNPs are preselected based on their association with the exposure trait (*P_GWAS_* < 1×10^−3^).

### Metabolites causally associated with psychiatric illness share genetic relationships with structural features of the cerebral cortex

Alteration of cerebral anatomy is a common feature of many psychiatric conditions [36, 37]. We therefore examined genetic relationships between metabolites causally associated with psychiatric conditions and measures of the cerebral surface area and thickness using GWAS from the ENIGMA consortium [38]. We specifically focussed on 12 metabolites with at least moderate evidence for partial genetic causality (LCV models), and the five HDL-related traits associated with AN (CAUSE; Fig. 4a). At an *FDR_BH_* < 0.05, four metabolites displayed consistent patterns of negative genetic correlation with global and region-specific cortical surface area (*r_g_* ≤ –0.08), with phospholipids to total lipids ratio in chylomicrons and extremely large VLDL most consistently represented (16 correlations; Fig. 4b,c, Table S10, S11). In contrast, 12 metabolites were positively correlated with surface area and thickness measures (*r_g_* ≥ 0.07), with average diameter for HDL particles most frequently correlated (19 correlations; Fig. 4b,c, Table S10, S11).

**Figure 4.**
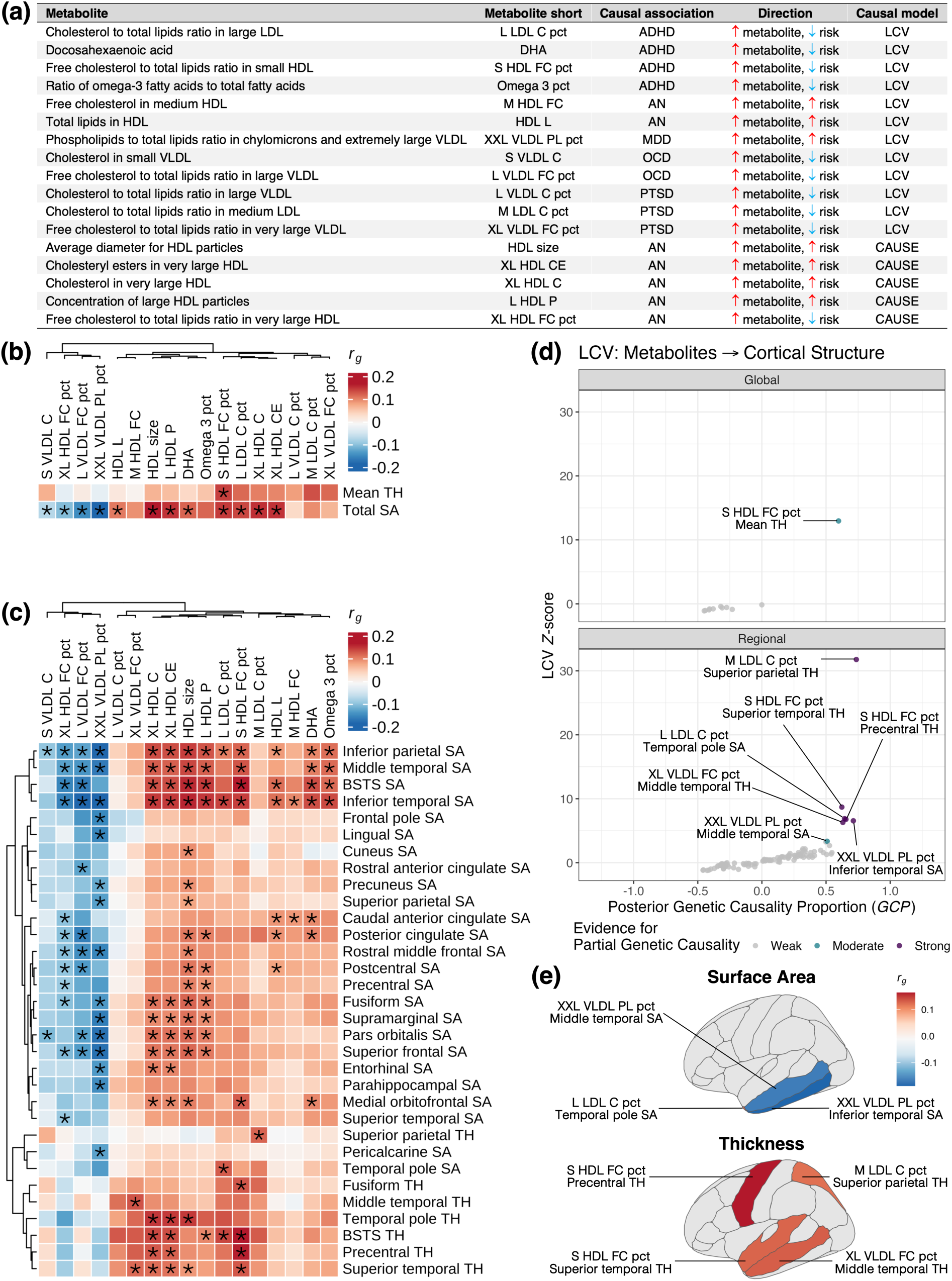
Genetic relationship between metabolites causally associated with psychiatric conditions and structure of the cerebral cortex. **(a)** Table summarising evidence for causal relationships between metabolites and psychiatric conditions. **(b, c)** Heatmaps presenting genetic correlation coefficients (*r_g_*) for the 17 metabolites reported in panel **(a)** with respect to mean cortical thickness (TH) and total surface area (SA) **(b)**, and region-specific measures of SA and TH **(c)**. The regional results have been subset to only include regions with at least one significant genetic correlation. * = *FDR_BH_* < 0.05. All columns and rows (regional measures only) have been subject to hierarchal clustering. **(d)** Scatter plots comparing LCV posterior genetic causality proportions (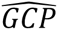) to their corresponding *Z*-scores. Purple points = trait pairings with strong evidence for partial genetic causality (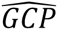 ≥ 0.6); teal points = trait pairings with moderate evidence (0.5 ≤ 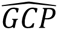 < 0.6). **(e)** Genetic correlation coefficients for trait pairings from panel **(d)** with strong or moderate evidence for causality, projected onto the left cerebral hemisphere (lateral view).

LCV models revealed strong evidence for partial genetic causality as follows: cholesterol to total lipids ratio in medium LDL → superior parietal thickness (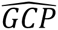 = 0.74), phospholipids to total lipids ratio in chylomicrons and extremely large VLDL → inferior parietal surface area (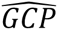 = 0.71), free cholesterol to total lipids ratio in small HDL → precentral thickness (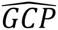 = 0.66), cholesterol to total lipids ratio in large LDL → temporal pole surface area (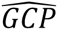 = 0.64), free cholesterol to total lipids ratio in very large VLDL → middle temporal thickness (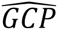 = 0.63), and free cholesterol to total lipids ratio in small HDL → superior temporal thickness (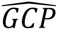 = 0.62; Fig. 4d, Table S12, S13). Moderate evidence was also uncovered for free cholesterol to total lipids ratio in small HDL → mean cortical thickness (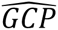 = 0.597) and phospholipids to total lipids ratio in chylomicrons and extremely large VLDL → middle temporal surface area (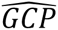 = 0.51; Fig. 4d, Table S12, S13). All relationships involving cortical thickness exhibited positive genetic correlation, suggesting elevation of these metabolites is associated with elevated thickness of these regions, whereas negative correlation was observed for surface area (Fig. 4c, 4e, Table S10, S11). One exception was cholesterol to total lipids ratio in large LDL → temporal pole surface area, which was positively correlated (Fig. 4c, 4e, Table S12, S13). While CAUSE models did not identify significant evidence for causality, these findings nonetheless suggest some metabolites causally associated with psychiatric conditions may also influence cortical structure (Table S14, S15).

We additionally examined genetic relationships between these cortical measures and psychiatric traits to explore evidence for causal chains, i.e.: metabolite → cortical measure → psychiatric trait. Significant genetic correlation was uncovered for inferior temporal surface area and MDD, (*r_g_* = –0.128, *SE* = 0.027, *P* = 3.27×10-6), middle temporal surface area and MDD, (*r_g_* = –0.087, *SE* = 0.028, *P* = 0.002), precentral thickness and ADHD, (*r_g_* = –0.138, *SE* = 0.047, *P* = 0.003) and middle temporal thickness and PTSD (*r_g_* = –0.109, *SE* = 0.044, *P* = 0.013) after correction for multiple testing (Table S16). However, no evidence for causality was uncovered (Table S17, S18).

### Some metabolites share gene-level common variant associations with psychiatric conditions

We next sought to identify specific genes that could mediate the interrelationship between metabolites and psychiatric conditions where stronger evidence was uncovered for shared genetic architecture than causality. The forwards CAUSE analysis initially revealed 137 metabolite-psychiatric trait pairings with greater evidence for horizontal pleiotropy than a causal relationship (i.e. *FDR_BH_* < 0.05, ΔELPD > 0; Fig. 3a, Table S6). Among these, 23 exhibited at least nominal evidence for horizontal pleiotropy in reverse models (*P* < 0.05, ΔELPD > 0), and were thus deemed strong candidates for further analysis (Fig. 5a, Table S19). For each trait pairing, SNPs were aggregated to genes using the Multimarker Analysis of GenoMic Annotation (MAGMA), after which common genes surpassing correction for multiple testing (*P_Bonf_* < 0.05) for both the metabolite and psychiatric condition were identified.

**Figure 5.**
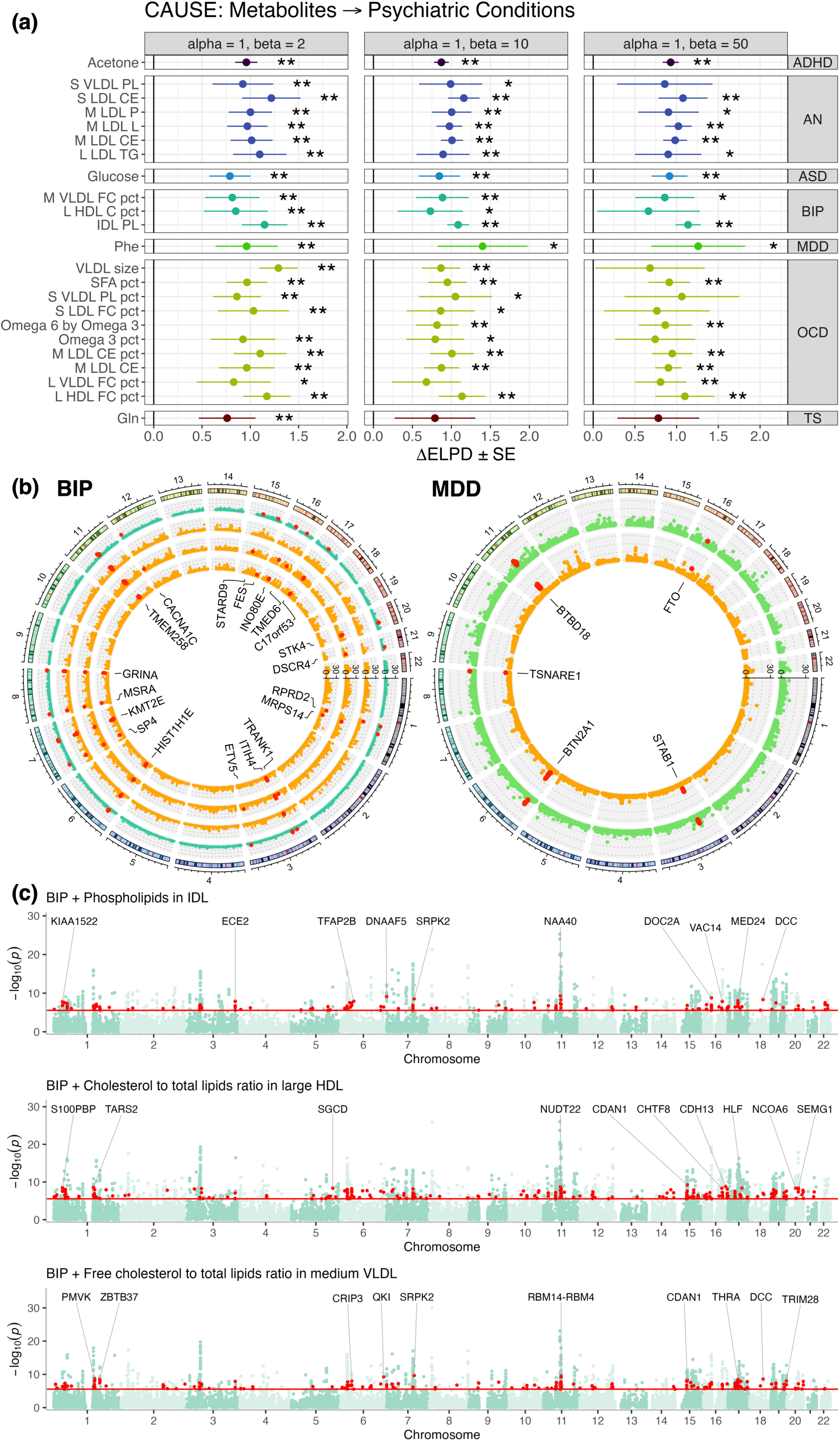
Shared gene-level common variant associations between metabolites and psychiatric illness. **(a)** Heatmap depicting *ΔELPD* estimates from the forwards CAUSE analysis (i.e. metabolite as exposure), subset to trait pairings that also exhibited nominal evidence for shared biology in the reverse analysis (i.e. psychiatric condition as exposure). * = *P* < 0.05, ** = *FDR_BH_* < 0.05. **(b)** Circos plots presenting shared gene-level common variant associations for BIP, MDD and their respective associated metabolites presented in panel **(a)**. Red points = genes with a Bonferroni-corrected *P_MAGMA_* < 2.6 × 10^−6^ for the psychiatric trait and at least one metabolite. Each shell of each plot represents a different trait as follows, radiating inwards: (left) BIP, phospholipids in IDL, cholesterol to total lipids ratio in large HDL and free cholesterol to total lipids ratio in medium VLDL; (right) MDD and phenylalanine. **(c)** Manhattan plots depicting pairwise MAGMA gene-level meta-analyses for BP and the same metabolites in panel **(b)**. Each point represents a single gene, with – log_10_(*P_meta_*) from the meta-analyses plotted on the y-axis. Red points = genes that were nominally significant when each trait was analysed separately but surpassed a Bonferroni correction for multiple testing (*P* < 2.6 × 10^−6^, horizontal red line) in the meta-analysis.

Interestingly, only BIP and MDD shared gene associations with their respective metabolites (Fig. 5b, Table S20). For BIP, 81 genes were shared with at least one of three corresponding metabolites, specifically: cholesterol to total lipids ratio in large HDL (65 genes), free cholesterol to total lipids ratio in medium VLDL (60) and Phospholipids in IDL (41; Fig. 5b, Table S20). Several shared genes were related to neuronal function, including ion channels and transporters (e.g. *CACNA1C*, *KCNS1*, *SLC12A9*, *SLC4A1*), regulators of synaptic function and neurotransmission (*ACHE*, *GLT8D1*, *GRINA*, *PACS1*, *TRANK1*, *TRIM38*), and transcription factors associated with neuronal development and plasticity (*ETV5*, *MYRF*, *RCOR2*, *SP4*), noting these functional associations are not exhaustive or mutually exclusive (Fig. 5b, Table S20). In addition, genes related to metabolic function (e.g. *COX8A, FADS1*, *FADS2*, *GAL3ST3*, *MSRA*), epigenetic regulation (including 10 histone proteins, *HDAC5*, *INO80E*, *KMT2E*, *PBRM1*) and immune function (*CTSF*, *FES*, *FEN1*, *ITIH1*, *ITIH3, ITIH4, STAB1*, TLR9, *TRIM38*, *WFDC5*) were also uncovered. A total of 37 genes were also shared between MDD and phenylalanine, including several that were identified in the analysis of BIP (e.g. *FADS1*, *FADS2*, *GLT8D1*, *ITIH1*, *MYRF*, *PBRM1, TRIM38*). Among genes exclusively shared between MDD and phenylalanine, we identified regulators of neuronal development and function (e.g. *BTBD18*, *CLP1*, *CTNND1*, *TSNARE1*, *ZDHHC5*) and immune function (*BTN2A1*, *BTN3A2*, *SERPING1*). Another notable shared association was the fat mass and obesity-associated gene *FTO*, a brain-enriched RNA N6-methyladenosine demethylase associated with energy homeostasis, satiety and obesity [39]. Despite these overlapping associations, no shared canonical pathways (Molecular Signatures Database, 3,917 pathways) were identified via a gene-set analysis after correction for multiple testing (*FDR_BH_* < 0.05).

### Pairwise gene and gene-set meta-analyses reveal novel associations between metabolites and psychiatric traits

Gene associations for all 23 trait pairings were meta-analysed to identify novel associations among genes that were nominally significant (*P_Bonf_* < *P* < 0.05) in the respective individual MAGMA analyses. Novel associations were uncovered for all 23 trait pairings, with BIP and cholesterol to total lipids ratio in large HDL exhibiting the greatest number of novel genes (*N_novel_* = 263, top hit = *CDAN1*, *P_meta_* = 5.74 × 10^−10^; Fig. 5c, Fig. S4, Table 2, Table S21, S22). BIP also returned the greatest number of novel associations per meta-analysis, with an average of 203 genes per metabolite (Table 2). Strikingly, genes related to neuronal development and function were consistently identified in association with BIP and all three corresponding metabolites, including *CACNB2*, *CDH13*, *DOC2A*, *HOMER2*, *NRCAM*, *S100PBP*, *SLC29A2*, *SNAP91* (Fig. 5d, Table S21, S22). Notably, the dopamine receptor *DRD2*, a gene commonly targeted by psychotropic medications, was also represented in the meta-analysis of BIP/free cholesterol to total lipids ratio in medium VLDL (Table S21, S22). Genes pertinent to neuronal function were also uncovered amongst the top hits for other trait pairings, including *SV2A* (ASD/glucose), *MCPH1* (OCD/Phospholipids to total lipids ratio in small VLDL), *KCND2* (OCD/ Ratio of omega-3 fatty acids to total fatty acids) and *RAB40C* (OCD/ Ratio of omega-6 fatty acids to omega-3 fatty acids; Table 2). Many of the top meta-analysis associations were also related to lipid, carbohydrate and amino acid metabolism, including: *PITPNM2* (ADHD/acetone), *LGR4* (AN /phospholipids in small VLDL), *ALDH18A1* (AN/total lipids in medium LDL), *SLC2A4* (AN /triglycerides in large LDL), *NAA40* (BIP/phospholipids in IDL), *ACO1* (MDD/ phenylalanine), *NDUFS3* (OCD/Cholesteryl esters to total lipids ratio in medium LDL) and *TRIT1* (OCD/Free cholesterol to total lipids ratio in large VLDL)

**Table 2.** Number of novel genic associations between metabolites and psychiatric conditions after MAGMA gene-level meta-analysis.

| Psychiatric condition | Metabolite | $N_{\text{novel associations}} (P_{\text{Bonf}} < 0.05)$ | Top gene | $P_{\text{meta}}$ |
| --- | --- | --- | --- | --- |
| **ADHD** | Acetone | 23 | *PITPNM2* | $9.93 \times 10^{-9}$ |
| **AN** | Cholesteryl esters in medium LDL | 35 | *ODF4* | $6.92 \times 10^{-8}$ |
| | Cholesteryl esters in small LDL | 47 | *ARHGEF15* | $1.64 \times 10^{-8}$ |
| | Concentration of medium LDL particles | 38 | *ARHGEF15* | $9.1 \times 10^{-8}$ |
| | Phospholipids in small VLDL | 50 | *LGR4* | $4.64 \times 10^{-8}$ |
| | Total lipids in medium LDL | 45 | *ALDH18A1* | $1.25 \times 10^{-7}$ |
| | Triglycerides in large LDL | 42 | *SLC2A4* | $5.67 \times 10^{-8}$ |
| **ASD** | Glucose | 17 | *SV2A* | $1.66 \times 10^{-7}$ |
| **BIP** | Cholesterol to total lipids ratio in large HDL | 263 | *CDAN1* | $5.74 \times 10^{-10}$ |
| | Free cholesterol to total lipids ratio in medium VLDL | 204 | *SRPK2* | $2.13 \times 10^{-10}$ |
| | Phospholipids in IDL | 143 | *NAA40* | $4.88 \times 10^{-10}$ |
| **MDD** | Phenylalanine | 70 | *ACO1* | $1.42 \times 10^{-9}$ |
| **OCD** | Average diameter for VLDL particles | 17 | *MTCH2* | $7.42 \times 10^{-7}$ |
| | Cholesteryl esters in medium LDL | 5 | *CORO1B* | $2 \times 10^{-6}$ |
| | Cholesteryl esters to total lipids ratio in medium LDL | 13 | *NDUFS3* | $6.23 \times 10^{-7}$ |
| | Free cholesterol to total lipids ratio in large HDL | 12 | *GAK* | $1.04 \times 10^{-6}$ |
| | Free cholesterol to total lipids ratio in large VLDL | 4 | *TRIT1* | $5.3 \times 10^{-7}$ |
| | Free cholesterol to total lipids ratio in small LDL | 16 | *KIT* | $2.11 \times 10^{-7}$ |
| | Phospholipids to total lipids ratio in small VLDL | 6 | *MCPH1* | $6.6 \times 10^{-7}$ |
| | Ratio of omega-3 fatty acids to total fatty acids | 8 | *KCND2* | $6.44 \times 10^{-7}$ |
| | Ratio of omega-6 fatty acids to omega-3 fatty acids | 10 | *RAB40C* | $8.08 \times 10^{-7}$ |
| | Ratio of saturated fatty acids to total fatty acids | 1 | *DRG2* | $5.84 \times 10^{-7}$ |
| **TS** | Glutamine | 24 | *RANBP10* | $5.44 \times 10^{-7}$ |

A similar meta-analysis was conducted for all associated pathways that were nominally significant in the individual MAGMA analyses. At an *FDR_BH_* < 0.05, 28 significant pathways were identified across 10 trait pairings, reflecting a broad range of biological functions. The meta-analysis of BIP and cholesterol to total lipids ratio in large HDL returned the largest number of gene-set associations (11 novel pathways), including the *MAPK pathway*, *Brain-derived neurotropic factor (BDNF) signalling*, *Insulin signalling* and *Serotonin receptor 4/6/7 and NR3C signalling*, among others (Table 3, Table S23, S24). This was followed by AN/triglycerides in large LDL, which shared: *Leptin pathway, Obesity pathway*, *Abacavir transmembrane transport* and *MET receptor activation* gene-sets, and ASD/glucose, which shared the *SMAD2/3 pathway*, *Hemostasis* and *Opioid receptor pathways* (Table 3, Table S23, S24). Overall, these results reveal novel gene-level common variant associations shared between psychiatric conditions and various metabolites, that in some cases converge on significant pathways.

**Table 3.**
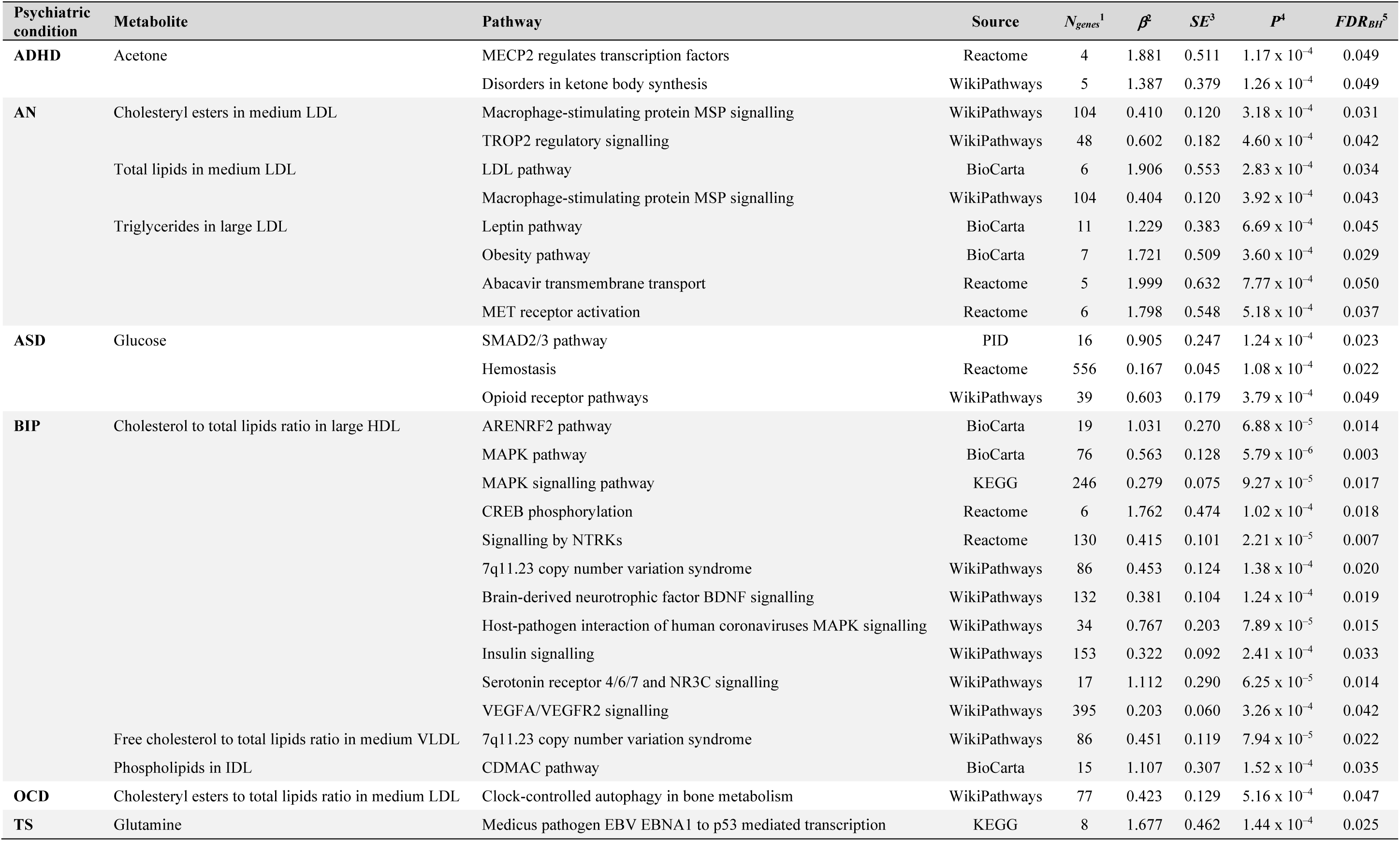
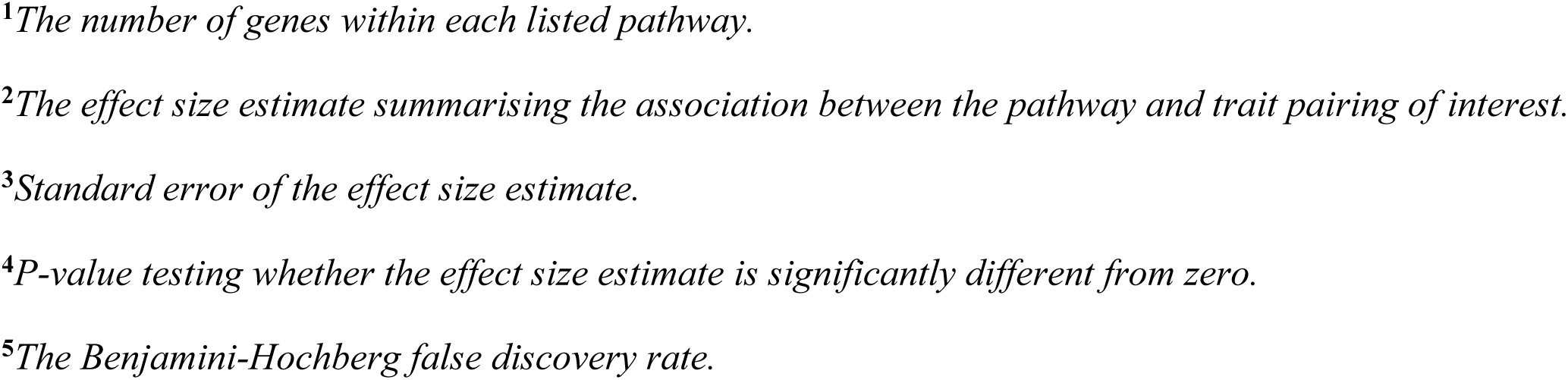
MAGMA pathway-level meta-analysis.

| Psychiatric condition | Metabolite | Pathway | Source | $N_{genes}^1$ | $\beta^2$ | $SE^3$ | $P^4$ | $FDR_{BH}^5$ |
| --- | --- | --- | --- | --- | --- | --- | --- | --- |
| **ADHD** | Acetone | MECP2 regulates transcription factors | Reactome | 4 | 1.881 | 0.511 | $1.17 \times 10^{-4}$ | 0.049 |
| | | Disorders in ketone body synthesis | WikiPathways | 5 | 1.387 | 0.379 | $1.26 \times 10^{-4}$ | 0.049 |
| **AN** | Cholesteryl esters in medium LDL | Macrophage-stimulating protein MSP signalling | WikiPathways | 104 | 0.410 | 0.120 | $3.18 \times 10^{-4}$ | 0.031 |
| | | TROP2 regulatory signalling | WikiPathways | 48 | 0.602 | 0.182 | $4.60 \times 10^{-4}$ | 0.042 |
| | Total lipids in medium LDL | LDL pathway | BioCarta | 6 | 1.906 | 0.553 | $2.83 \times 10^{-4}$ | 0.034 |
| | | Macrophage-stimulating protein MSP signalling | WikiPathways | 104 | 0.404 | 0.120 | $3.92 \times 10^{-4}$ | 0.043 |
| | Triglycerides in large LDL | Leptin pathway | BioCarta | 11 | 1.229 | 0.383 | $6.69 \times 10^{-4}$ | 0.045 |
| | | Obesity pathway | BioCarta | 7 | 1.721 | 0.509 | $3.60 \times 10^{-4}$ | 0.029 |
| | | Abacavir transmembrane transport | Reactome | 5 | 1.999 | 0.632 | $7.77 \times 10^{-4}$ | 0.050 |
| | | MET receptor activation | Reactome | 6 | 1.798 | 0.548 | $5.18 \times 10^{-4}$ | 0.037 |
| **ASD** | Glucose | SMAD2/3 pathway | PID | 16 | 0.905 | 0.247 | $1.24 \times 10^{-4}$ | 0.023 |
| | | Hemostasis | Reactome | 556 | 0.167 | 0.045 | $1.08 \times 10^{-4}$ | 0.022 |
| | | Opioid receptor pathways | WikiPathways | 39 | 0.603 | 0.179 | $3.79 \times 10^{-4}$ | 0.049 |
| **BIP** | Cholesterol to total lipids ratio in large HDL | ARENRF2 pathway | BioCarta | 19 | 1.031 | 0.270 | $6.88 \times 10^{-5}$ | 0.014 |
| | | MAPK pathway | BioCarta | 76 | 0.563 | 0.128 | $5.79 \times 10^{-6}$ | 0.003 |
| | | MAPK signalling pathway | KEGG | 246 | 0.279 | 0.075 | $9.27 \times 10^{-5}$ | 0.017 |
| | | CREB phosphorylation | Reactome | 6 | 1.762 | 0.474 | $1.02 \times 10^{-4}$ | 0.018 |
| | | Signalling by NTRKs | Reactome | 130 | 0.415 | 0.101 | $2.21 \times 10^{-5}$ | 0.007 |
| | | 7q11.23 copy number variation syndrome | WikiPathways | 86 | 0.453 | 0.124 | $1.38 \times 10^{-4}$ | 0.020 |
| | | Brain-derived neurotrophic factor BDNF signalling | WikiPathways | 132 | 0.381 | 0.104 | $1.24 \times 10^{-4}$ | 0.019 |
| | | Host-pathogen interaction of human coronaviruses MAPK signalling | WikiPathways | 34 | 0.767 | 0.203 | $7.89 \times 10^{-5}$ | 0.015 |
| | | Insulin signalling | WikiPathways | 153 | 0.322 | 0.092 | $2.41 \times 10^{-4}$ | 0.033 |
| | | Serotonin receptor 4/6/7 and NR3C signalling | WikiPathways | 17 | 1.112 | 0.290 | $6.25 \times 10^{-5}$ | 0.014 |
| | | VEGFA/VEGFR2 signalling | WikiPathways | 395 | 0.203 | 0.060 | $3.26 \times 10^{-4}$ | 0.042 |
| | Free cholesterol to total lipids ratio in medium VLDL | 7q11.23 copy number variation syndrome | WikiPathways | 86 | 0.451 | 0.119 | $7.94 \times 10^{-5}$ | 0.022 |
| | Phospholipids in IDL | CDMAC pathway | BioCarta | 15 | 1.107 | 0.307 | $1.52 \times 10^{-4}$ | 0.035 |
| **OCD** | Cholesteryl esters to total lipids ratio in medium LDL | Clock-controlled autophagy in bone metabolism | WikiPathways | 77 | 0.423 | 0.129 | $5.16 \times 10^{-4}$ | 0.047 |
| **TS** | Glutamine | Medicus pathogen EBV EBNA1 to p53 mediated transcription | KEGG | 8 | 1.677 | 0.462 | $1.44 \times 10^{-4}$ | 0.025 |

<sup>1</sup>*The number of genes within each listed pathway.*
<sup>2</sup>*The effect size estimate summarising the association between the pathway and trait pairing of interest.*
<sup>3</sup>*Standard error of the effect size estimate.*
<sup>4</sup>*P-value testing whether the effect size estimate is significantly different from zero.*
<sup>5</sup>*The Benjamini-Hochberg false discovery rate.*

## DISCUSSION

Metabolites related to cardiometabolic health are often dysregulated in psychiatric conditions, yet it is unclear if these metabolites are causally associated with psychiatric health or share biology relevant to clinical management. In this study, we systematically analysed genetic correlation and causation between circulating metabolites and psychiatric conditions to explore putative causal relationships and identify biologically significant genes shared between these traits. Broad patterns of genetic correlation were uncovered between lipid traits and psychiatric illness, with strong representation from fatty acid, cholesterol and lipoprotein traits, amongst others. This is consistent with observational and genetic evidence indicating that dyslipidaemia is a common feature of many psychiatric conditions, wherein alteration of lipid profiles is thought to impact variables such as inflammation, neuronal structure and neurotransmission [27, 28, 40–43]. Similarly, amino-acids (e.g. glutamine and tyrosine) and glycolysis-related traits (e.g. glucose and citrate) have been previously associated with psychiatric health [10, 44–48]. In the case of glycaemic traits, it is well established that severe mental illnesses such as schizophrenia exhibit a strong comorbidity with type-2 diabetes irrespective of treatment with psychotropic medications [49, 50]. Increasing evidence also suggests conditions such as MDD and SZ are associated with dysregulation of amino acids, particularly those associated with neurotransmission (e.g. glutamine, tryptophan and tyrosine) [51–53]. Despite this, we note that many genetically correlated metabolites did not show evidence for a causal relationship with psychiatric illness in the present study. This conflicts with recent Mendelian randomisation studies that have reported evidence for causality for some of these metabolites, including polyunsaturated fatty acids, triglycerides and glucose [41, 44, 54]. Disparity between these findings may reflect methodological differences between conventional MR approaches and the causal models employed in this study. For example, the CAUSE MR framework models correlated pleiotropy and other confounders that conventional MR methods fail to address, thus CAUSE may be more robust to false positives [32]. We also utilised the largest uniformly processed metabolite GWAS to date [25], which may provide the statistical power required to clarify previously reported causal associations. Nonetheless, the pleiotropic effect of these variants on other traits could also mediate these relationships, therefore, these metabolites may nonetheless offer promising biomarkers and opportunities for clinical intervention in psychiatric illness.

There was consistent evidence that elevation of HDL-related traits is causally associated with increased odds of anorexia nervosa. Robust evidence was identified in relation to the average diameter and concentration of HDL particles, and cholesterol/cholesteryl ester content of large/very large HDL particles. Our findings therefore suggest these specific features of HDL are most pertinent to AN risk, building upon previous observational studies reporting elevated blood HDL concentrations amongst individuals with AN [55, 56], as well as genetic studies reporting positive genetic correlation between these variables [57]. We consider our findings to be particularly robust since evidence for causality was retained after removing variants associated with BMI, which represents a potential confounder given that low BMI is both associated with altered lipoprotein profiles and required for AN diagnosis [34]. However, the biological mechanism through which these HDL-related traits could impact AN risk remains unclear. To explore causal effects associated with the brain, we examined whether features of HDL are causally associated with structural properties of the cerebral cortex, wherein many disruptions associated with AN have been previously reported [58–60]. Although no strong evidence for causality was uncovered, genetic correlation was identified between HDL-related traits and structural features of regions previously associated with AN, including the superior frontal, insula and precuneus regions [58–60], among others, necessitating further analysis is to deconvolute any potential association between HDL and AN mediated through brain alterations. Finally, we acknowledge that free cholesterol content of very large HDL was associated with decreased odds of AN, indicating further exploration is required to ascertain this direction of effect and its biological significance.

LCV models also revealed evidence for a causal, risk-increasing relationship for phospholipid ratios in chylomicrons and extremely large VLDL on MDD. Previous observational studies have consistently reported dyslipidaemia as a feature of MDD [40, 61], with recent genetic evidence further suggesting a causal role for specific circulating lipids, including triglycerides and HDL [41]. However, there is limited evidence pertaining a relationship with phospholipids, chylomicrons and/or VLDL. A recent metabolome-wide association study of MDD using UK Biobank participants revealed no association with phospholipid ratios in chylomicrons and extremely large VLDL, and this was further supported by Mendelian randomisation [62]. Although some observational studies have identified decreased ether phospholipids [63] and VLDL [64] among individuals with depressive symptoms, this conflicts with the direction of effect reported in the current study, suggesting further analysis is required to deconvolute these results. We also uncovered evidence suggesting elevated LDL and/or VLDL cholesterol content is protective for OCD and PTSD. Notably, few studies have examined these specific traits in either condition. In the case of PTSD, observational studies have consistently reported elevation of total cholesterol and LDL among individuals with this condition [65, 66], however it is unclear whether the cholesterol content of LDL is specifically altered. There is also limited evidence pertaining dysregulation of VLDL in OCD, with some observational evidence suggesting serum VLDL is elevated in OCD [67], while other studies have reported little-to-no association [68]. We therefore emphasise the need for further investigation of LDL-/VLDL-related traits in OCD and PTSD to clarify the association between these variables, dissect correlation from causation, and identify specific features of LDL/VLDL with potential clinical utility in these conditions.

In addition to these traits, LCV models uncovered evidence suggesting elevated docosahexaenoic acid, omega-3 fatty acid to total fatty acid ratios, free cholesterol in small HDL and cholesterol in large LDL are associated with decreased odds of ADHD. Previous observational studies have shown that docosahexaenoic acid, and omega-3 fatty acids more generally, are decreased amongst individuals with ADHD (versus healthy controls) and correlate with cognitive symptoms [69, 70]. Strikingly, a meta-analysis of randomised controlled trials has further shown that dietary supplementation of these compounds significantly improves clinical symptoms of children and adolescents with ADHD [71]. In the case of LDL and HDL, there is conflicting evidence pertaining an association with ADHD, with some studies suggesting these traits are altered in the serum of affected individuals [72, 73], while others report limited association [74, 75]. We nonetheless believe these lipoprotein traits represents a key area for further investigation since both exhibited evidence for partial genetic causality with respect to structural features of cortical brain regions previously associated with ADHD [76–79]. Specifically, these traits were associated with elevated precentral and superior temporal thickness and temporal pole surface area. To this end, multivariable causal modelling would be useful to further explore whether the putative protective effect of these lipid traits in ADHD is elicited via alteration of cortical structure [80].

Extensive overlap of gene-level common variant associations was uncovered between 23 pairings of metabolites and psychiatric conditions, thus providing candidate genes that could mediate the interrelationship between these traits in the absence of causal relationships. We consider these findings to be particularly robust given that all 23 trait pairings were genetically correlated and exhibited evidence for horizontal pleiotropy in forwards and reverse CAUSE models. Strong overlap of gene-level common variant associations was identified between BIP and lipoprotein-related traits, converging on genes and/or pathways related to neuronal development and function, metabolic processes, epigenetic regulation and immune function, amongst others. In some cases, these genes provide additional functional significance to those previously associated with BIP. For instance, *FADS1/2* have been recently implicated in a putative causal relationship between BIP and arachidonic acid [54] and prioritised for drug repurposing [8], with our results further suggesting these genes may also link BIP and lipoprotein-related traits. Similarly, genes associated with synaptic function (e.g. *CACNB2*, *HOMER2*, *NRCAM*, and *SNAP91*) that may contribute to neural circuit dysregulation in BIP [81] were also related to lipoprotein traits, emphasising the potential functional significance of this horizontal pleiotropy. In this case, it is possible that lipoprotein traits are interrelated with biological processes such as synaptic vesicle release that are also dysregulated in BIP [81]. Future work should also explore whether relationships such as these converge on gene networks associated with psychotropic medications, which are known to elicit a range of metabolic side effects [82]. In the present study, we identified shared common variant signatures at the *DRD2* gene between BIP and free cholesterol to total lipids ratio in medium VLDL. This is particularly noteworthy given that DRD2 is a target of many approved antipsychotic medications [83], and considering our results, may also be interrelated with properties of VLDL. This is further supported by recent work revealing DRD2 expressing neurons can be modulated by lipoprotein lipase (LPL), which itself is responsive to dietary triglycerides [84]. Another interesting finding from this analysis was the overlap of gene-associations between AN and LDL-/VLDL-related traits. This stands in contrast to the causal relationships observed for HDL-associated traits on AN, suggesting the presence of a particularly complex relationship between circulating lipoproteins and AN risk. Collectively, our results further support the existence of gene-level horizontal pleiotropy between some psychiatric conditions and specific metabolites that requires deeper exploration to further dissect the significance of this shared biology.

In summary, our results present evidence for putative causal relationships and shared genetic architecture between circulating metabolites and psychiatric conditions with potential utility for clinical management. There are some limitations that need to be considered when interpreting these results. Firstly, the analyses conducted in this study are subject to inherent limitations of the GWAS data, such as population stratification [85] and other biases. For example, we note that the UK Biobank is composed of individuals over the age of 40, thus this age bias may have affected GWAS for the metabolic traits [86]. Similarly, our analyses were conducted using data from individuals of predominantly European ancestry, limiting the applicability of our findings to other ethnic superpopulations. All trait pairings with evidence for a causal relationship also require further validation via randomised controlled trials. We nonetheless attempted to replicate our findings using GWAS from [26], however we note that results for the CAUSE analysis failed to yield consistent results, potentially owing to the substantially reduced sample size of these GWAS. We nonetheless believe the CAUSE methodology offers a strong framework for exploring putative causal relationships between complex polygenic traits in future work, as it mitigates false positives associated with horizontal pleiotropy and other potential confounders [32]. Finally, our exploration of brain regions causally associated with specific metabolites was not exhaustive, and future work should include additional brain structures (e.g. subcortical structures), and measures (e.g. connectivity) that are also related to psychiatric illness [87]. In summary, we believe the current study offers a strong platform for using GWAS to guide prioritisation of specific metabolites for future study in psychiatric illness in relation to causal mechanisms, shared biology and precision medicine.

## METHODS

### Study overview

The primary aim of this work was to better understand how clinically actionable metabolites impact psychiatric health by systematically exploring the genetic relationship between these traits using SNPs (Fig. 1). We specifically leveraged the largest, uniformly processed metabolite GWAS available to explore genetic correlation, followed by genetic causal inference using Latent Causal Variable (LCV) models and the Causal Analysis Using Summary Effect estimates (CAUSE) methodology. The goal of this analysis was to uncover metabolites with evidence for a causal effect on psychiatric conditions and therefore generate a resource that prioritises these metabolites for further investigation via clinical trials. In addition, the CAUSE methodology was also used to identify trait pairings with stronger evidence for shared genetic architecture, rather than a causal relationship. These traits were specifically analysed for shared, gene level common variant signatures using the Multimarker Analysis of GenoMic Annotation (MAGMA), with a view of both identifying common genes and mechanisms that may mediate the interplay between metabolic and psychiatric health.

### GWAS summary statistics

GWAS summary statistics for 10 psychiatric conditions were obtained from the Psychiatric Genomics Consortium (PGC), prioritising the largest available GWAS of predominantly European ancestry. We specifically examined attention deficit hyperactivity disorder (ADHD, *N* = 225,534, [15]), anorexia nervosa (AN, *N* = 72,517, [16]), autism spectrum disorders (ASD, *N* = 46,351, [17]), bipolar disorder (BIP, *N* = 840,309, [18]), major depressive disorder (MDD, *N* = 2,000,702, [19]), obsessive compulsive disorder (OCD, *N* = 9,725, [20]), panic disorder (PD, *N* = 9,907, [21]), post-traumatic stress disorder (PTSD, *N* = 1,249,840, [22]), schizophrenia (SZ, *N* = 130,644, [23]) and Tourette syndrome (TS, *N* = 14,307, [24]).

GWAS summary statistics for 249 metabolites were obtained from the large, uniformly processed meta-analysis (*N* = 599,529) of 185,352 individuals from the Estonian Biobank and 413,897 individuals of European ancestry from the UK Biobank, conducted by [25]. The metabolites include triglycerides, fatty acids, phospholipids, cholesterol, ketone bodies, glycolysis related metabolites and amino acids, amongst others. All metabolites were measured in EDTA plasma samples via nuclear magnetic resonance (NMR) and inverse normal transformed prior to within-cohort association testing, using the sex, age, age^2^ and genetic principal components (top 10 for EstBB, top 20 for UKB) as covariates. For all metabolite-psychiatric trait pairings with significant evidence for causality, sensitivity analyses were conducted using an independent metabolite GWAS from [26], a meta-analysis of 136,016 individuals across 33 independent (non-Estonian or UK Biobank) cohorts. GWAS of structural measures of the cerebral cortex were obtained from the ENIGMA consortium [38]. We specifically used GWAS of global and regional surface area and thickness, generated from a meta-analysis of 33,992 individuals of predominantly European ancestry across 49 cohorts. All data were covaried for age, age^2^, sex, sex-by-age and age^2^ interactions, the first four genetic multidimensional scaling components, diagnostic status and scanner. For all summary statistics, we excluded 1) palindromic variants, 2) non-SNP variants (e.g. insertions, deletions), 3) variants with low imputation accuracy, where available (INFO < 0.9).

### Genetic correlation

Genetic correlation was estimated between all metabolites and psychiatric conditions via linkage disequilibrium score regression (LDSR), as implemented in the *ldsc* package (v1.0.1) [29, 30]. Briefly, LDSR estimates genetic covariance between traits by regressing SNP level *χ*^2^ values – the product of SNP *Z*-scores from each trait – against LD scores that estimate total LD for a given SNP. Genetic covariance is then normalised to trait heritabilities to obtain genetic correlation (*r_g_*), noting that an intercept term is included that mitigates confounding due to factors such as sample overlap between GWAS.

All summary statistics were firstly harmonised into a “munged” format, wherein SNP effect sizes and standard errors were transformed into *Z*-scores, ensuring the sign of each *Z*-score was relative to the statistical effect allele. We also retained approximately 1 million high-confidence HapMap3 SNPs outside the major histocompatibility complex region (MHC, chr6:28000000– 34000000) with minor allele frequency (MAF) > 0.05. Genetic correlation was then estimated using the *ldsc.py* script, with LD scores sourced from the 1000 Genomes Project European reference panel (available at: https://alkesgroup.broadinstitute.org/LDSCORE/). Trait pairings surpassing a Benjamini-Hochberg false discovery rate (*FDR_BH_*) < 0.05 were considered significant and retained for causal inference.

### Latent causal variable modelling

Evidence for causality was assessed amongst genetically correlated trait pairings using a Latent Causal Variable model (LCV, [31]). LCV assumes a latent variable, *L*, mediates the genetic correlation between two traits. As such, if trait 1 has stronger genetic correlation with *L* than trait 2, trait 1 is deemed to be partially genetically causal for trait 2. Partial genetic causality is quantified as a posterior genetic causality proportion (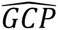), derived by comparing the mixed fourth moments (co-kurtosis) of SNP marginal effect size distributions between two traits. This leverages the fact that if trait 1 is partially genetically causal for trait 2, most SNPs affecting trait 1 will have proportional effects on trait 2 but not vice versa. 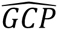 estimates between 0 (no genetic causality) and ±1 (full genetic causality) summarise the strength of evidence for a causal relationship for trait 1 on trait 2 and strictly do not serve as a magnitude of causal effect. Positive 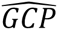 values indicate evidence that trait 1 is partially genetically causal for trait 2, whereas negative values suggest trait 2 exhibits evidence for causality on trait 1. As shown previously, trait pairings with 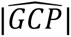 estimates ≥ 0.6, directionally consistent *Z*-scores and *FDRbh* < 0.05 were considered to exhibit strong evidence for partial genetic causality [31]. Trait pairings fulfilling these criteria with 0.5 ≤ 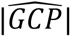 < 0.6 were deemed to have moderate evidence for partial genetic causality. All summary statistics were munged prior to analysis as recommended [88].

### CAUSE

In addition to LCV, the Causal Analysis Using Summary Effect estimates (CAUSE) method (v1.2.0.0335, [32]) was also used to estimate causal relationships amongst genetically correlated traits. Briefly, CAUSE is a method of Mendelian randomisation (MR) that uses SNPs associated with an exposure (i.e. metabolites) as instrumental variables (IVs) to explore evidence for causality with respect to an outcome trait (i.e. psychiatric conditions). CAUSE specifically uses a Bayesian framework that estimates causal effects conditioned on confounding pleiotropy (i.e. IVs acting directly on the outcome or via a confounding variable) by employing a beta prior distribution. This approach is applied to independent SNPs that are broadly associated with the exposure (*P*_GWAS_ < 1 × 10^−3^) to simultaneously capture causal relationships, correlated pleiotropy and uncorrelated pleiotropy. The ability to condition on correlated pleiotropy is particularly advantageous, as this cannot be readily assessed using conventional tests for pleiotropy (e.g. Cochran’s *Q* test, Egger intercept). Unlike many other summary-based MR models, CAUSE also includes a correlation term that empirically estimates and adjusts for sample overlap between the exposure and outcome and other confounders.

For all genetically correlated trait pairings, we used CAUSE to specifically compare models of genetic causality with horizontal pleiotropy against sharing models that only examine horizontal pleiotropy by fixing the causal effect (γ) at zero. These models were assessed using three distinct beta prior distributions, which assume high (⍺ = 1, β = 2), moderate (⍺ = 1, β = 10, default prior) or low correlated pleiotropy (⍺ = 1, β = 50), the former representing the most conservative but least powered model, while the latter is the most well-powered (and similar to traditional summary-based MR), but can be susceptible to false positives. Model comparisons were expressed as a change in expected log pointwise posterior density (*ΔELPD*), wherein negative values suggest the causal model fits better than sharing the sharing model, while positive values suggest the sharing model provides the best fit. This was examined for trait pairings with Pareto *k* estimates < 0.67, whereas those with Pareto *k* ≥ 0.67 were excluded from further analyses. Metabolite-psychiatric trait pairings with significant evidence for causality were defined as those with a negative *ΔELPD* that surpassed an *FDR_BH_* < 0.05, and also exhibited no evidence for causality in reverse models that used the psychiatric trait as the exposure and metabolite as outcome. We also define trait pairings with significant evidence for shared genetic architecture as those with a positive *ΔELPD* that surpassed an *FDR_BH_* < 0.05 in the forwards analysis, and *P* < 0.05 in the reverse analyses.

### Gene and gene-set association analysis

Common variant signatures for metabolites or psychiatric conditions were aggregated within protein coding genes using the Multimarker Analysis of GenoMic Annotation (MAGMA, v1.10, Linux, [33]). This analysis was restricted to all trait pairings with evidence for shared biology as nominated by CAUSE, with the aim of comparing shared gene and gene-set associations between each nominated trait pairing. GWAS SNPs were firstly mapped to 19,240 autosomal protein-coding genes in the GRCh37 genome assembly (NCBI), available at: https://vu.data.surfsara.nl/index.php/s/Pj2orwuF2JYyKxq. Genic boundaries were extended to 5kb upstream and 1.5kb downstream to include potential regulatory variants, and furthermore, we excluded genes within the MHC region due to the complexity of LD in this region. Gene-level association was then assessed via the default gene-based test in MAGMA, which uses a linear combination of SNP-level GWAS *P-*values to produce mean *χ^2^* test-statistics for each gene. To account for dependent *P*-values due to LD between variants, the 1000 genomes phase 3 European reference panel was used to derive variant-level LD, which weight the contribution of each SNP to the final association test-statistic. Gene associations with a Bonferroni-corrected *P* < 2.6 × 10^−6^ were deemed statistically significant, adjusting for 19,240 independent tests for each phenotype.

The MAGMA competitive gene-set association test was then employed to identify canonical pathways from the Molecular Signatures Database (MSigDB, 3917 pathways) associated with each phenotype of interest. MAGMA specifically constructs linear regression models that test whether genic association (transformed to *Z*-scores via the probit function) is stronger within the gene set of interest, compared to all other genes, with covariates for gene size and gene minor allele count. Gene-set associations with *FDRbh* < 0.05 were deemed significant.

### Pairwise gene and gene-set meta-analysis

Genic *Z*-scores for metabolites and psychiatric traits with evidence for shared biology were subjected to pairwise meta-analyses using the --meta flag in MAGMA. This utilises Stouffer’s weighted *Z* method, which combines *Z*-scores for each gene (*i*) weighted by the GWAS sample size (*ω_i_*) as follows:

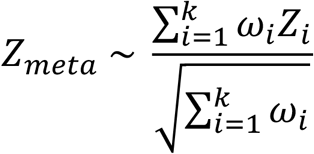

To mitigate bias due to sample overlap and other confounders, the LDSR intercept for the metabolite and psychiatric condition of interest was included as a covariate as recommended by the MAGMA authors. Gene-set meta-analyses were additionally undertaken using the model described above, with *Z_meta_* as the outcome variable in each instance.

## ACKNOWLEDGEMENTS

This work was supported by National Health and Medical Research Council (NHMRC) grants (G1147644, G1188493). M.J.C. is supported be an NHMRC Senior Research Fellowship (G1121474). W.R.R. is supported by an NHMRC Investigator Fellowship (EL1, 2025671). The funders had no role in study design, data collection, analysis, decision to publish or preparation of the manuscript.

## DATA AVAILABILITY

All GWAS summary statistics utilised in this study are publicly available and can be accessed via the Psychiatric Genomics Consortium website (https://pgc.unc.edu/for-researchers/download-results/) or the GWAS Catalog with accession numbers GCST90451106–GCST90451354 [25], and GCST90301941–GCST90302173 [26]. Sample scripts used in the present study can be accessed at: https://github.com/D-Kiltschewskij/Genetic_Correlation_Causation_Metabolic_Psychiatric_Traits.

## AUTHOR CONTRIBUTIONS

D.J.K. designed the study, curated and processed the data, conducted and interpreted the statistical analyses and wrote the manuscript. W.R.R. assisted with study design and interpretation of the results, provided methodological insights and edited the manuscript. M.J.C. provided funding, contributed to interpretation of the results and edited the manuscript.

