## Supplementary Figures for "A Genetic Atlas of Relationships Between Circulating Metabolites and Liability to Psychiatric Conditions"

**Contents**

**Page 2–4.	Figure S1.** Genetic correlation among psychiatric conditions and traits related to lipoprotein subclasses.

**Page 5.	Figure S2.** Reverse CAUSE models examining the effect of psychiatric traits on metabolites.

**Page 6.	Figure S3.** CAUSE posterior gamma estimates for HDL-related traits on AN, with BMI-associated SNPs excluded.

**Page 7–12.	Figure S4.** Manhattan plots for all MAGMA gene-level meta-analyses.

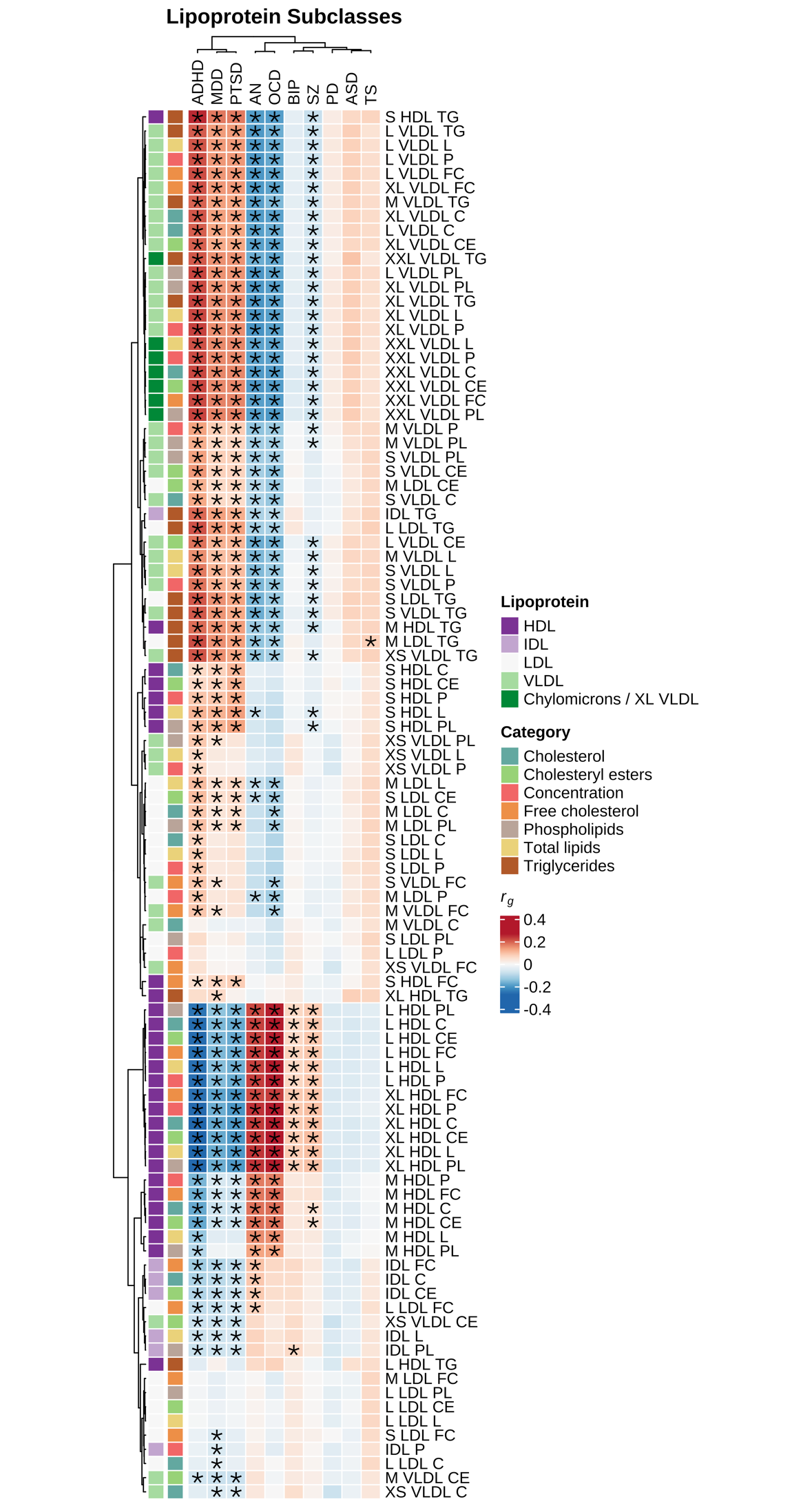

**(a)**

**Lipoprotein absolute lipid content**

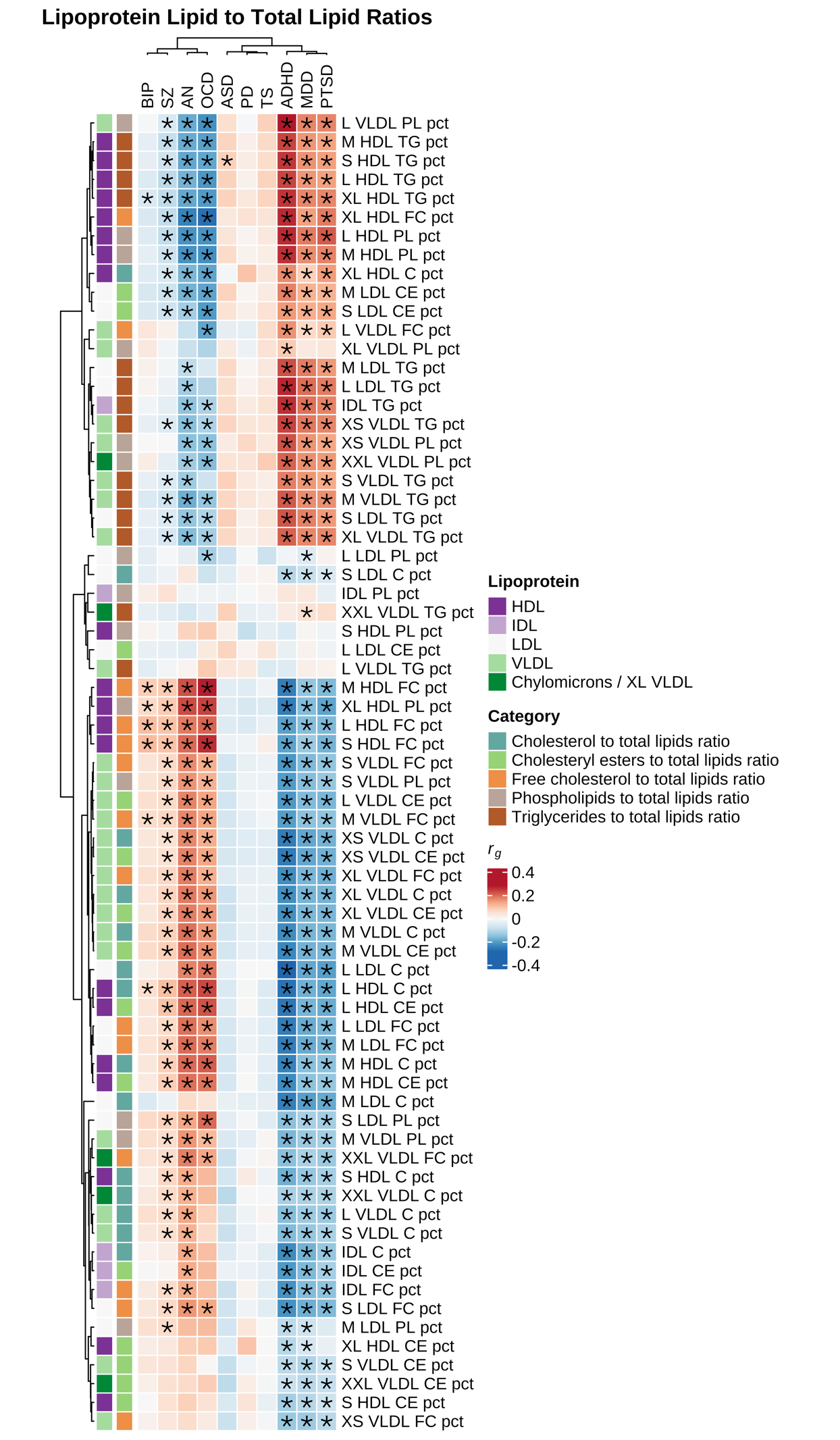

**(b)**

**Lipoprotein lipid ratios**

**Figure S1. Genetic correlation among psychiatric conditions and traits related to lipoprotein subclasses.** Heatmaps depicting LDSR genetic correlation coefficients (*r_g_*) between the 10 psychiatric conditions, **(a)** lipoprotein absolute lipid content and **(b)** lipoprotein lipid ratios, subset by lipoprotein particle diameter. Rows and columns were subject to hierarchal clustering to identify similar groups of traits. **FDR_BH_* < 0.05. Full metabolite names can be accessed in Table S1.

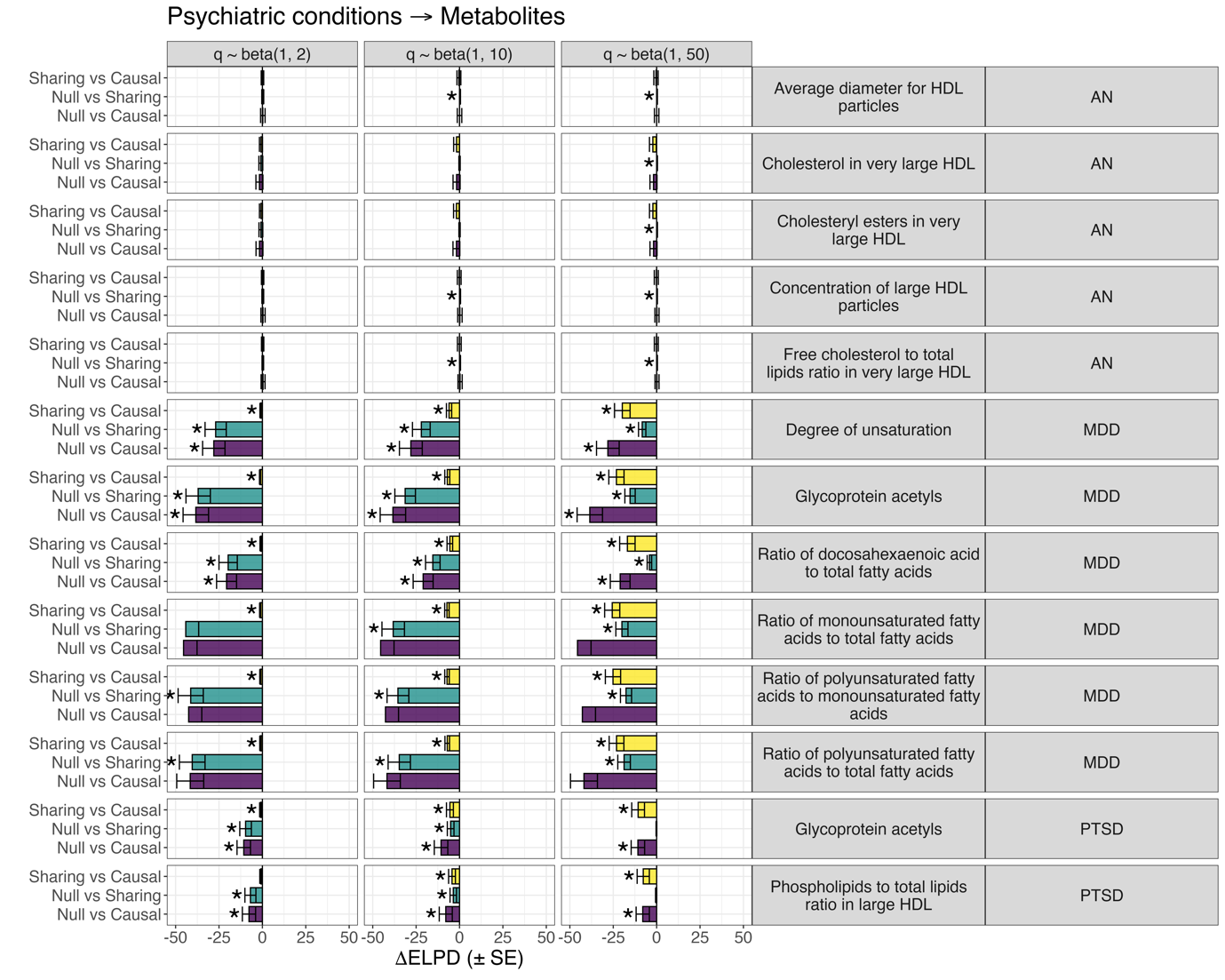

**Figure S2. Reverse CAUSE models examining the effect of psychiatric traits on metabolites.** *ΔELPD* (± standard error [*SE*]) estimates for CAUSE models examining the effect of psychiatric traits on metabolites, subset to trait pairings with evidence for causality in the forwards analysis. Yellow bars = the sharing versus causal model comparisons; teal bars = comparison of casual and null models; purple bars = comparison of sharing and null models. Note that a negative *ΔELPD* estimate indicates that the second model (as indicated on the left y-axis) fits better than the first. * = *P* < 0.05.

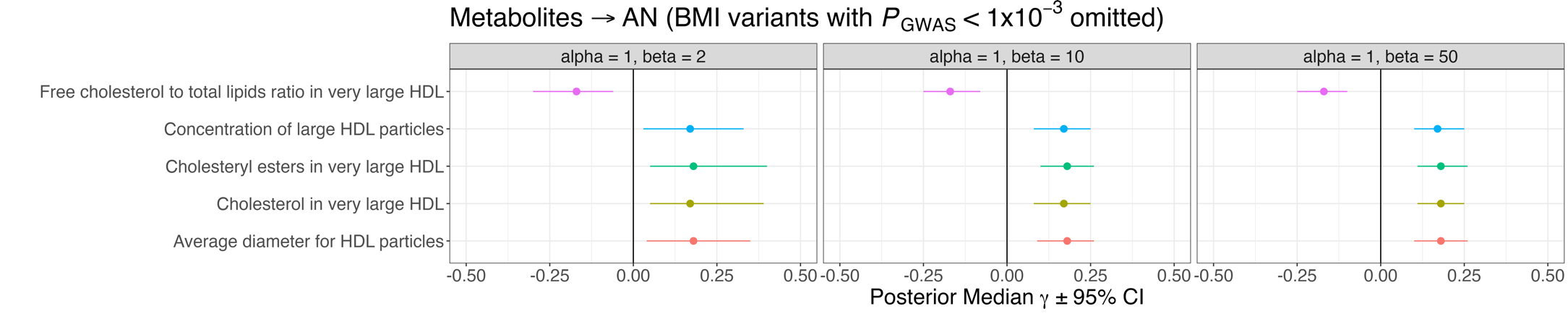

**Figure S3**. **CAUSE posterior gamma estimates for HDL-related traits on AN, with BMI-associated SNPs excluded.** Causal effect estimates (posterior median γ ± 95% credible interval [*CI*]) summarising direction of effect between all five HDL-related traits causally associated with AN, with BMI-associated variants excluded.

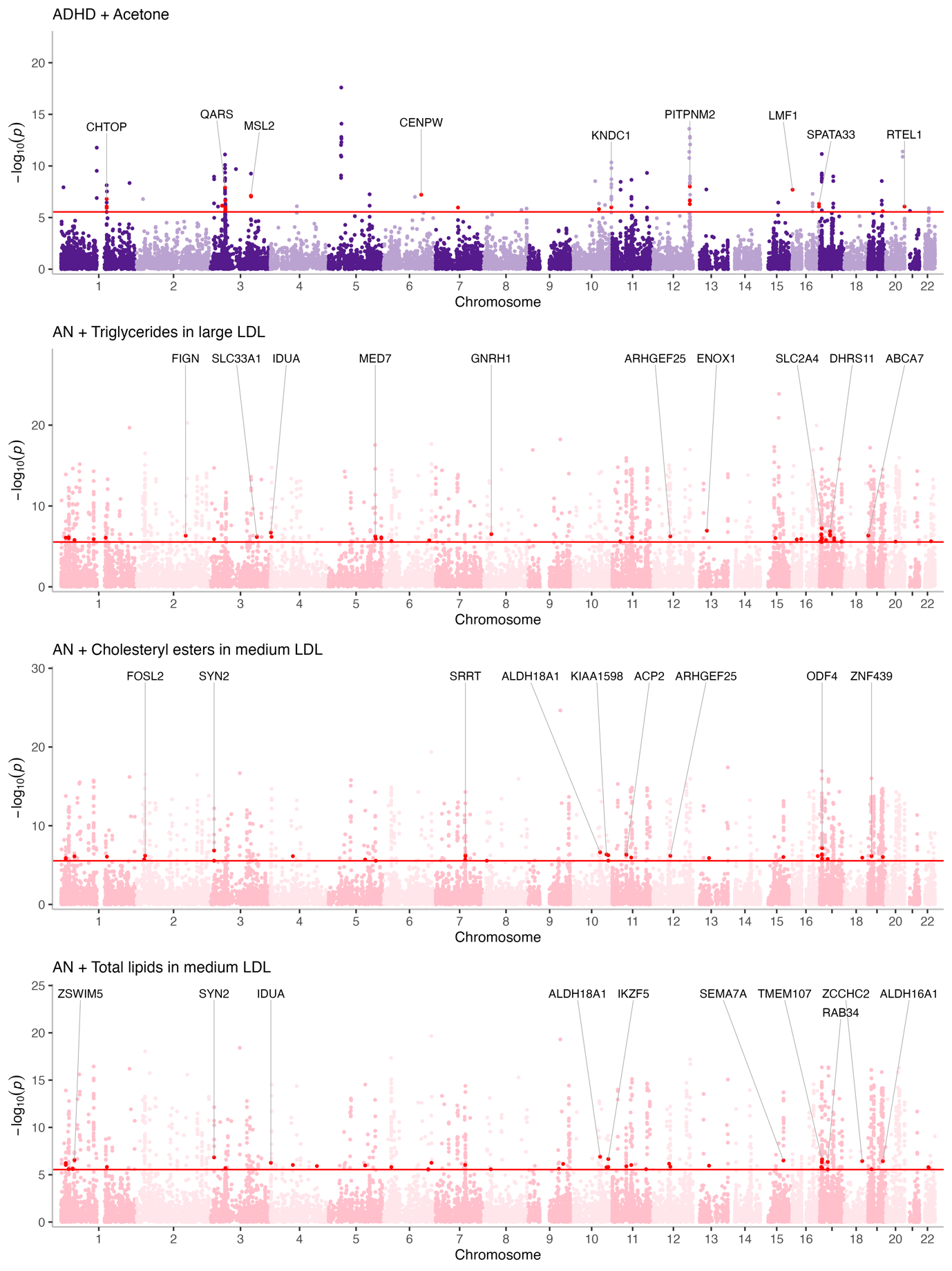

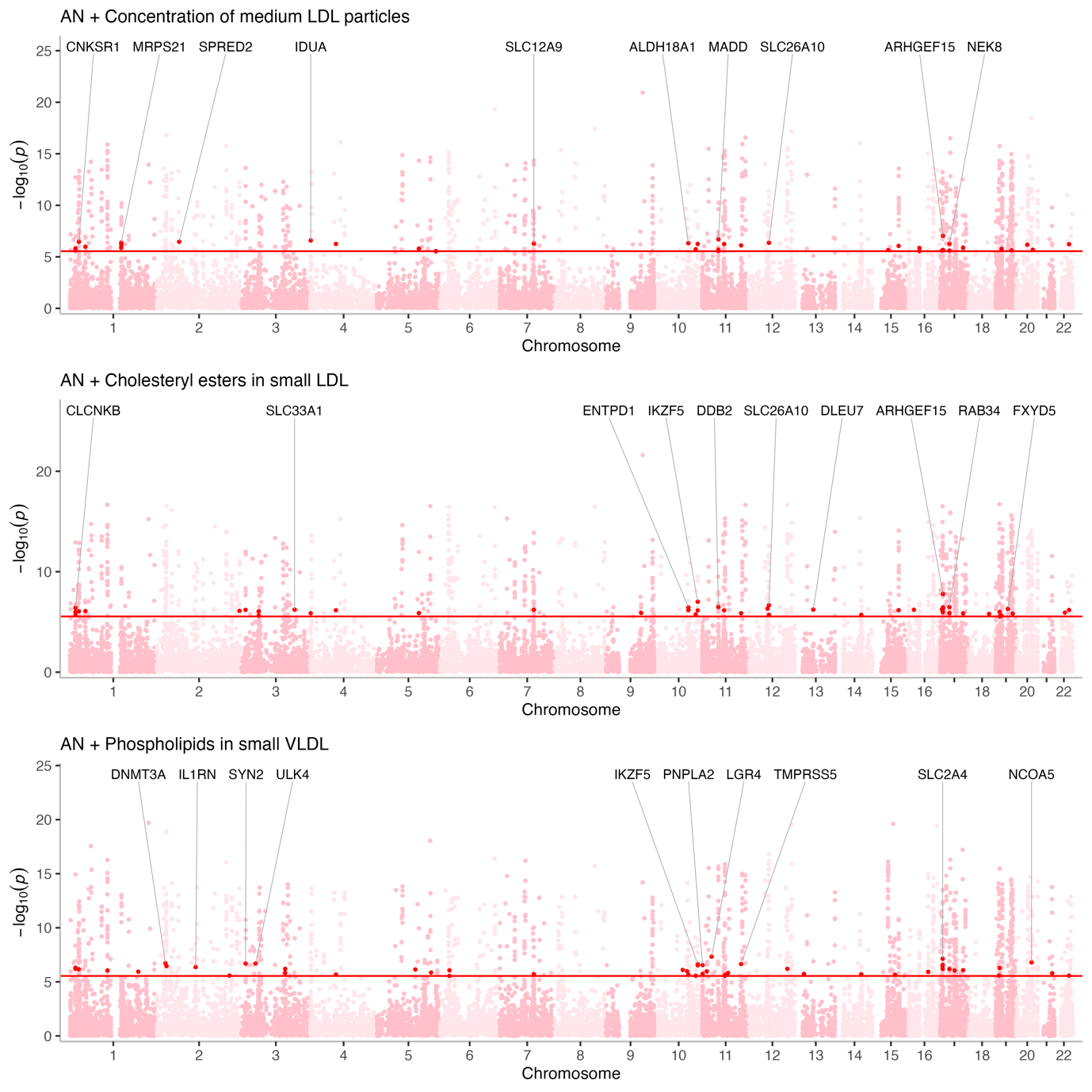

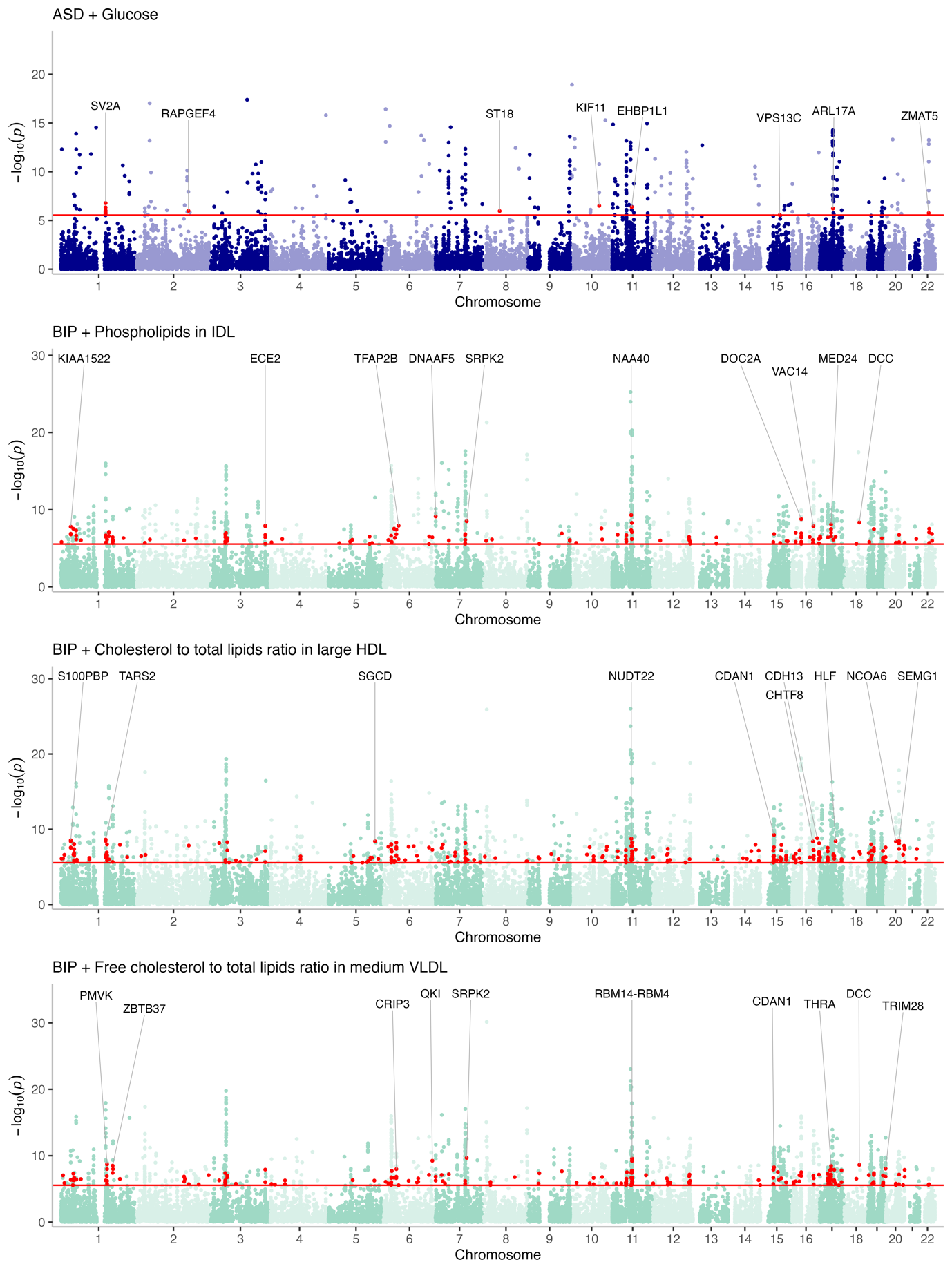

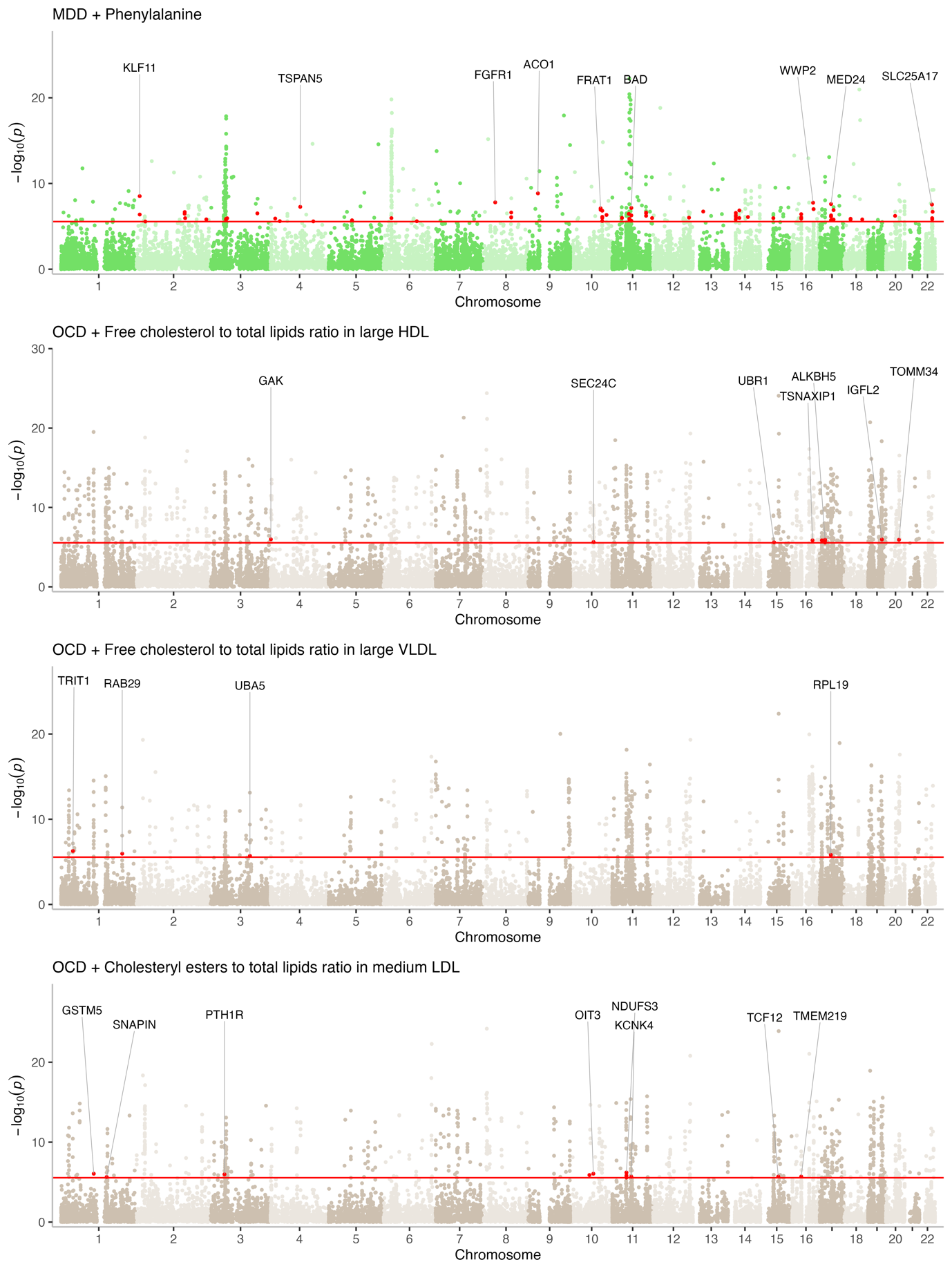

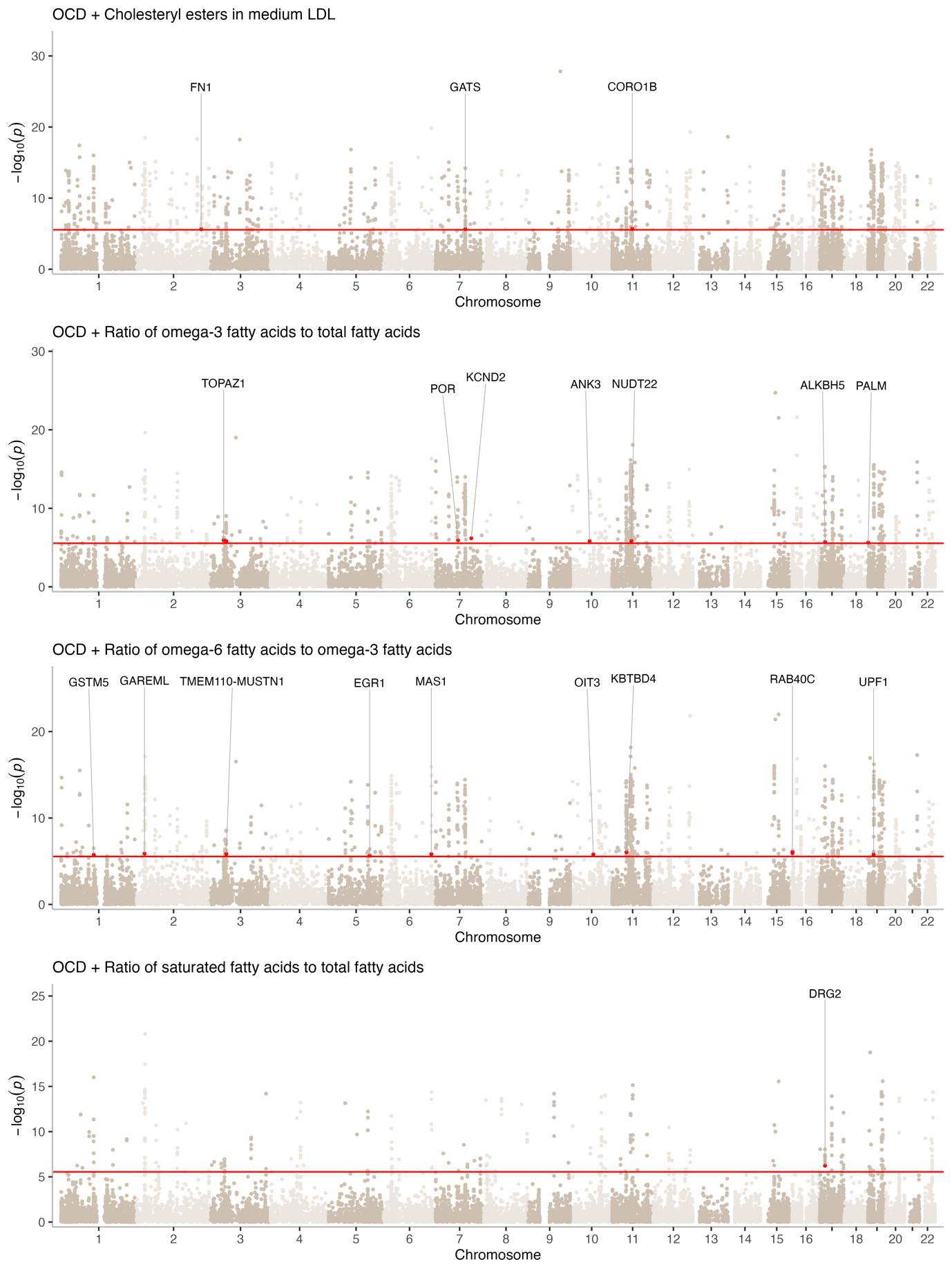

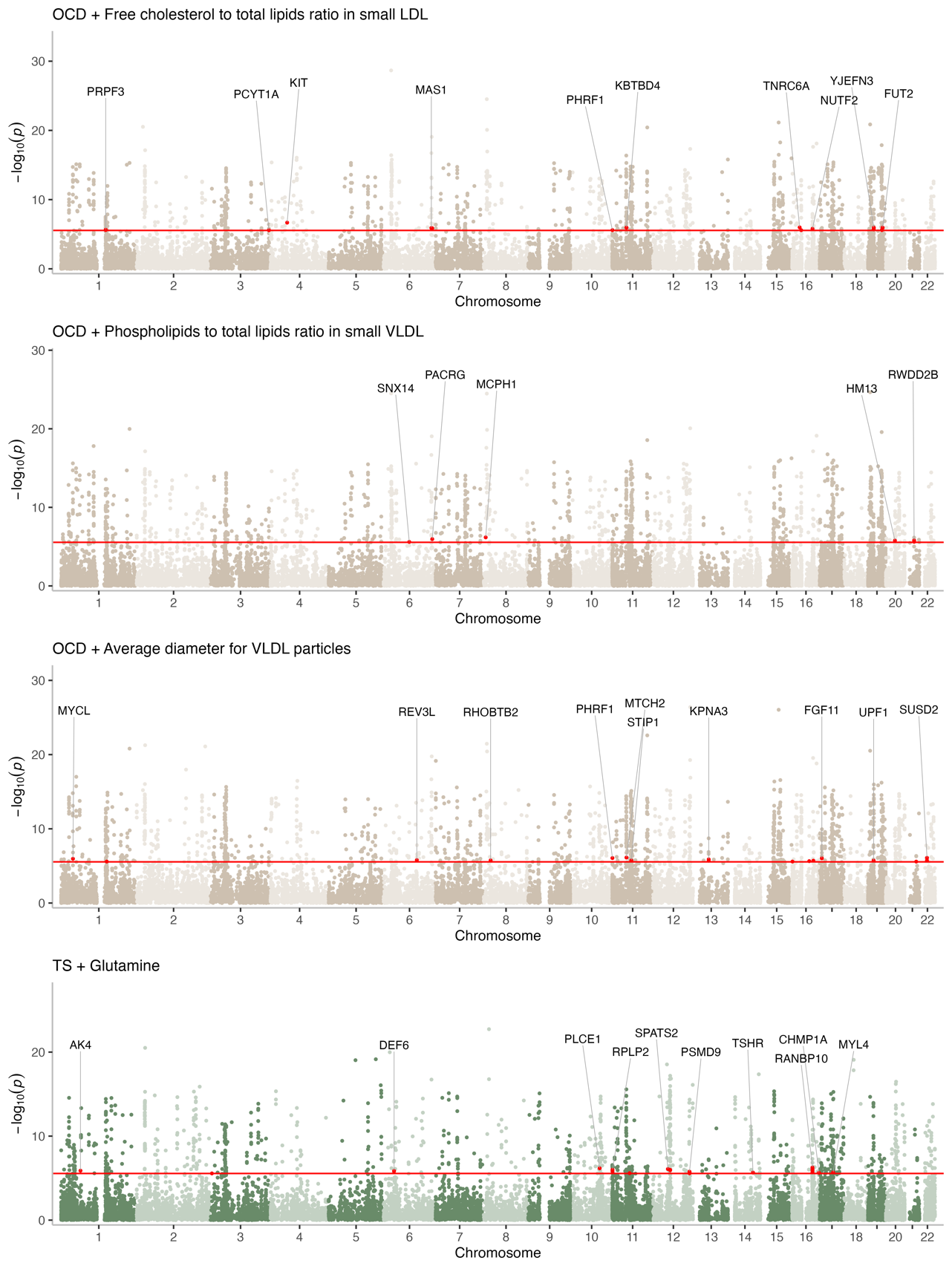

**Figure S4. Manhattan plots for all MAGMA gene-level meta-analyses.** Manhattan plots depicting pairwise MAGMA gene-level meta-analyses for all 23 metabolite-psychiatric condition pairings identified in Fig. 5a. Each point represents a single gene, with –log_10_(*P_meta_*) from the meta-analyses plotted on the y-axis. Red points = genes that were nominally significant when each trait was analysed separately but surpassed a Bonferroni correction for multiple testing (*P* < 2.6 x 10^–6^, horizontal red line) in the meta-analysis.
